# The transmission blocking activity of artemisinin-combination, non-artemisinin, and 8-aminoquinoline antimalarial therapies: a pooled analysis of individual participant data

**DOI:** 10.1101/2024.09.27.24314479

**Authors:** Leen N Vanheer, Jordache Ramjith, Almahamoudou Mahamar, Merel J Smit, Kjerstin Lanke, Michelle E Roh, Koualy Sanogo, Youssouf Sinaba, Sidi M. Niambele, Makonon Diallo, Seydina O Maguiraga, Sekouba Keita, Siaka Samake, Ahamadou Youssouf PharmD, Halimatou Diawara, Sekou F. Traore, Roly Gosling, Joelle M Brown, Chris Drakeley, Alassane Dicko, Will Stone, Teun Bousema

## Abstract

**Background:** Interrupting human-to-mosquito transmission is important for malaria elimination strategies as it can reduce infection burden in communities and slow the spread of drug resistance. Antimalarial medications differ in their efficacy in clearing the transmission stages of *Plasmodium falciparum* (gametocytes) and in preventing mosquito infection. Here we present a combined analysis of six trials conducted at the same study site with highly consistent methodologies that allows for a direct comparison of the gametocytocidal and transmission-blocking activities of fifteen different antimalarial regimens or dosing schedules.

**Methods and findings:** Between January 2013 and January 2023, six clinical trials with transmission endpoints were conducted at the Clinical Research Centre of the Malaria Research and Training Centre of the University of Bamako in Mali. These trials tested Artemisinin-Combination Therapies (ACTs), non-ACT regimens and combinations with 8-aminoquinolines. Participants were males and non-pregnant females, between 5-50 years of age, who presented with P. falciparum mono-infection and gametocyte carriage by microscopy. Blood samples were taken before and after treatment for thick film microscopy, infectivity assessments by mosquito feeding assays and molecular quantification of gametocytes. Mixed-effects generalized linear models were fit with individual-specific random effects and fixed effects for time points, treatment groups and their interaction. Models quantified changes in mosquito infection rates and gametocyte densities within treatment arms over time and between treatments. In a pooled analysis of 422 participants, we observed substantial differences between ACTs in gametocytocidal and transmission-blocking activities, with artemether-lumefantrine (AL) being significantly more potent at reducing mosquito infection rates within 48 hours than dihydroartemisinin-piperaquine (DHA-PPQ), artesunate-amodiaquine (AS-AQ) and pyronaridine-artesunate (PY-AS) (p<0.0001). The addition of single low dose primaquine (SLD PQ) accelerated gametocyte clearance and led to a significantly greater reduction in mosquito infection rate within 48-hours of treatment for each ACT, while an SLD of the 8-aminoaquinoline tafenoquine (TQ) showed a delayed but effective response compared to SLD primaquine. Finally, our findings confirmed considerably higher post-treatment transmission after sulfadoxine-pyrimethamine plus amodiaquine (SP-AQ) compared to most ACTs, with a significantly lower relative reduction in mosquito infection rate at day 7 compared to DHA-PPQ, AS-AQ, and AL (p<0.0001). Therefore, adding an SLD PQ to SP-AQ may be beneficial to block malaria transmission in community treatment campaigns.

**Conclusions:** We found marked differences among ACTs and single low-dose 8-aminoquinoline drugs in their ability and speed to block transmission. The findings from this analysis can support treatment policy decisions for malaria elimination and be integrated into mathematical models to improve the accuracy of predictions regarding community transmission and the spread of drug resistance under varying treatment guidelines.

## INTRODUCTION

The primary aim in the therapeutic management of malaria is the clearance of pathogenic asexual blood-stage parasites, using antimalarials with schizonticidal properties. Artemisinin-based combination therapies (ACTs) are the first-line treatment for uncomplicated *Plasmodium falciparum* malaria across the world and are highly potent against asexual parasites, capable of reducing the circulating asexual parasite biomass ∼10,000 fold per 48 hour cycle (1). Gametocytes are distinct parasite stages that are not associated with symptoms but are the only parasite life stage that can be transmitted to and establish infection in mosquitoes. In *P. falciparum*, gametocytes develop during five developmental stages over a prolonged 10–12-day maturation period. Importantly, *P. falciparum* gametocytes have markedly different sensitivity profiles to antimalarial drugs than asexual parasites. Assessing and comparing the gametocytocidal and transmission-blocking properties of different antimalarial regimens is important for informing treatment guidelines that aim to contribute to transmission reduction.

Artemisinins are active against developing gametocytes but less so against the mature forms (2). Although gametocyte density generally decreases more after treatment with ACTs compared to non-ACTs (3), there is considerable variation in gametocyte-clearing potential between ACTs (4). For example, current artemisinin partner drugs such as piperaquine, lumefantrine and amodiaquine have limited activity against *P. falciparum* gametocytes at clinically relevant concentrations (2,5–7). *In vitro* data for the partner drug pyronaridine are contradictory; one study found a strong effect against mature gametocytes (8), whereas others only observed activity against mature gametocytes at concentrations above the therapeutic threshold (2,6,9). Furthermore, a disconnect exists between the detection of gametocytes and infectivity to mosquitoes; on the one hand, gametocytes can be infectious at sub-microscopic densities (10), whereas on the other, antimalarials can have a parasite-inhibiting effect, active after ingestion by mosquitoes (2,11) or sterilise gametocytes so that these are detectable but not transmissible (12). Gametocyte quantification is therefore an imperfect measure of post-treatment transmission potential, and mosquito feeding assays are considered more informative (13).

With the threat of artemisinin partial resistance in sub-Saharan Africa (14), various strategies have been proposed to counter the spread of resistant parasites. One of the most promising approaches, according to modelling simulations, is the use of Triple Artemisinin-based Combination Therapies (TACT), that combine existing ACTs with a second partner drug. This approach may offer protection against partner drug resistance and maintain high treatment efficacy in areas where resistance against artemisinins and one of the partner drugs is present (15). Other suggested strategies include multiple first-line ACT therapies or cycling between different ACTs (16,17). An alternative or complementary strategy is to supplement treatment with gametocytocidal compounds. Primaquine (PQ) and its long-lasting analogue tafenoquine (TQ) are 8-aminoquinolines that can clear *P. vivax* liver stages (18) and have *P. falciparum* gametocytocidal activity. The WHO recommends supplementing first-line ACTs with a single low-dose (0.25mg/kg) PQ (SLD PQ) in low transmission areas (19), and the WHO malaria policy and advisory group has now suggested expanding this recommendation to other regions (20). TQ holds the promise of long-lasting transmission blocking activity and recently a single-low dose of TQ (SLD TQ) in combination with ACTs and non-ACT treatments was found to effectively block transmission within 7 days at a dose of 1.66 mg/kg (21,22). To date, no trials directly comparing SLD TQ to SLD PQ have been conducted, largely due to the impracticality of trials involving mosquito feeding assays.

In order to make informed decisions regarding optimal antimalarial regimens, it is necessary to understand and compare the transmission-blocking abilities of different ACTs and TACTs. We conducted a pooled analysis of individual-level data from six clinical trials to compare the transmission-blocking effects of 15 different antimalarials and combinations. All six trials measured mosquito transmission endpoints, used sensitive gametocyte quantification, and were conducted using the same mosquito feeding protocols and assays in Ouélessébougou, Mali, between 2013 and 2023 (21–26). Results from this analysis can be used in mathematical models to more accurately predict community transmission and the spread of drug resistance under different treatment guidelines, and will inform malaria control programmes.

## METHODS

### Study design and participants

Between January 2013 and January 2023, six clinical trials (21–26) (Table 1) (appendix 2) involving a total of 521 participants were performed at the Clinical Research Centre of the Malaria Research and Training Centre (MRTC) of the University of Bamako (Bamako, Mali). Study participants were recruited in the commune of Ouélessébougou, which includes the central town of Ouélessébougou and 44 surrounding villages, with an estimated 50,000 inhabitants and located approximately 80 km south of Bamako, the capital of Mali. Malaria transmission in this region is hyperendemic and highly seasonal with incidence peaking during and following the rainy season from July to November. In all six trials, participants were males and non-pregnant females, between 5-50 years of age, with a body weight less than 80 kg, who presented with *P. falciparum* mono-infection and at least 1 (study acronyms PQ03, NECTAR 1-4) or 2 (study PQ01) gametocytes per 500 white blood cells (WBC) detected by blood smear. This minimum gametocyte density corresponds to an approximate minimum of 16-32 gametocytes per µL of blood when assuming 8000 WBC per µL. Participants in all studies, except for the 2013-2014 PQ01 study, were exclusively asymptomatic, and four studies (study acronyms PQ01, PQ03, NECTAR2, NECTAR3) required participants to have normal glucose-6-phosphate dehydrogenase (G6PD) production, as determined by OSMMR-D-D calorimetric test (R&D Diagnostics, Aghia Paraskevi, Greece; studies PQ01, NECTAR2 and NECTAR3), CareStart G6PD rapid diagnostic test (Access Bio, Somerset, NJ,USA; study PQ03) and/or STANDARD G6PD quantitative enzyme activity test (SD Biosensor, Suwon, South Korea; studies NECTAR2 and NECTAR3; table 1, appendix 2). Other study-specific inclusion criteria are presented in appendix 2.

**Table 1.** Overview of the included studies. Rows in grey indicate treatment groups that were not included in the analyses.

| Study | Year | Treatment group | Number of participants | Treatment category | Study population | Reference |
| --- | --- | --- | --- | --- | --- | --- |
| PQ01 | 2013-2014 | DHA-PPQ | 16 | DHA-PPQ | G6PD+ males, 5-50 years, $\geq 32$ gametocytes/μL | (24) |
|  |  | DHA-PPQ + 0.0625 mg/kg PQ | 16 | / |  |  |
|  |  | DHA-PPQ + 0.125 mg/kg PQ | 17 | / |  |  |
|  |  | DHA-PPQ + 0.25 mg/kg PQ | 15 | ACT-PQ |  |  |
|  |  | DHA-PPQ + 0.5 mg/kg PQ | 17 | ACT-PQ |  |  |
| PQ03 | 2016 | SP-AQ | 20 | SP-AQ | G6PD+ males, 5-50 years, asymptomatic, $\geq 16$ gametocytes/μL | (23) |
|  |  | SP-AQ + 0.25 mg/kg PQ | 20 | Non-ACT-PQ |  |  |
|  |  | DHA-PPQ | 20 | DHA-PPQ |  |  |
|  |  | DHA-PPQ + MB | 20 | / |  |  |
| NECTAR1 | 2019 | DHA-PPQ | 25 | DHA-PPQ | Males and non-pregnant females, 5-50 years, asymptomatic, $\geq 16$ gametocytes/μL | (25) |
|  |  | DHA-PPQ + 0.25 mg/kg PQ | 25 | ACT-PQ |  |  |
|  |  | PY-AS | 25 | PY-AS |  |  |
|  |  | PY-AS + 0.25 mg/kg PQ | 25 | ACT-PQ |  |  |
| NECTAR2 | 2020 | DHA-PPQ | 20 | DHA-PPQ | G6PD+ males and non-pregnant females, 12-50 years, asymptomatic, $\geq 16$ gametocytes/μL | (22) |
|  |  | DHA-PPQ + 0.42 mg/kg TQ | 20 | / |  |  |
|  |  | DHA-PPQ + 0.83 mg/kg TQ | 20 | ACT-TQ |  |  |
|  |  | DHA-PPQ + 1.66 mg/kg TQ | 20 | ACT-TQ |  |  |
| NECTAR3 | 2021 | AL | 20 | AL | G6PD+ males and non-pregnant females, 10-50 years, asymptomatic, $\geq 16$ gametocytes/μL | (21) |
|  |  | AL + 0.25 mg/kg PQ | 20 | ACT-PQ |  |  |
|  |  | SP-AQ | 20 | SP-AQ |  |  |
|  |  | SP-AQ + 1.66 mg/kg TQ | 20 | / |  |  |
| NECTAR4 | 2022 | AL | 20 | AL | Males and non-pregnant females, 10-50 years, asymptomatic, $\geq 16$ gametocytes/μL | (26) |
|  |  | AL-AQ | 20 | AL |  |  |
|  |  | AL-AQ + 0.25 mg/kg PQ | 20 | ACT-PQ |  |  |
|  |  | AS-AQ | 20 | AS-AQ |  |  |
|  |  | AS-AQ + 0.25 mg/kg PQ | 20 | ACT-PQ |  |  |

Ethical approvals for the individual studies were obtained from the Ethics Committee of the Faculty of Medicine, Pharmacy, and Dentistry of the University of Science, Techniques, and Technologies of Bamako (Bamako, Mali). In addition, the studies were approved by the Committee on Human Research at the University of California (San Francisco, CA, USA), and/or the Research Ethics Committee of the London School of Hygiene and Tropical Medicine (London, UK).

### Procedures

Of the antimalarial treatments evaluated in the included studies, the following were included in this analysis: i) Dihydroartemisinin-piperaquine (DHA-PPQ; Eurartesim; Sigma Tau, Gaithersburg, MD, USA); ii) Pyronaridine-artesunate (PY-AS; Pyramax; Shin Poong Pharmaceutical, Seoul, South Korea; iii) Artemether-lumefantrine (AL; Coartem; Novartis, Basel, Switzerland or Guilin Pharmaceutical, Shanghai, China); iv) Artesunate-amodiaquine (AS-AQ; Guilin Pharmaceutical, Shanghai, China); v) Sulfadoxine-pyrimethamine plus amodiaquine (SP-AQ; Guilin Pharmaceutical, Shanghai, China); vi) Any of the ACTs listed above plus a single low-dose of primaquine (ACT-PQ; 0.25 or 0.5 mg/kg; Sanofi, Laval, QC, Canada or ACE Pharmaceuticals, Zeewolde, the Netherlands); vii) Non-ACT (SP-AQ) plus a single low-dose of primaquine (Non-ACT-PQ; Sanofi, Laval, QC, Canada); viii) ACT (DHA-PPQ) plus a single low dose of tafenoquine (ACT-TQ; 0.83 or 1.66 mg/kg; 60° Pharmaceuticals Ltd, USA) (Table 1). Antimalarial treatments were administered as per manufacturer’s instructions (supplementary Information 1 for dosing tables and supplementary Table 1 for manufacturers, appendix 1, pp 3-6) under direct supervision. ACT treatments and sulfadoxine-pyrimethamine plus amodiaquine were administered over 3 days (days 0, 1, and 2). Primaquine and tafenoquine were administered as a single dose immediately after the first dose of ACT. In the NECTAR1 study, participants were treated with a full course of DHA-PPQ at day 21 of follow-up, to prevent re-infection. For the current analysis, we did not consider study arms within the included studies that are currently not considered as treatment regimens, such as arms with PQ doses below 0.25 mg/kg (n=33 participants) and with TQ doses below 0.83 mg/kg (n=20). Likewise, treatment arms involving methylene blue (n=20) or the combination of SP-AQ with an SLD TQ (n=20) were not considered in the analysis.

In all studies, blood samples were taken before treatment and on days 2 and 7 following treatment for thick film microscopy, infectivity assessments and molecular analysis of gametocyte density, as outlined in Figure 1 and described in detail in appendix 2.

**Figure 1.**
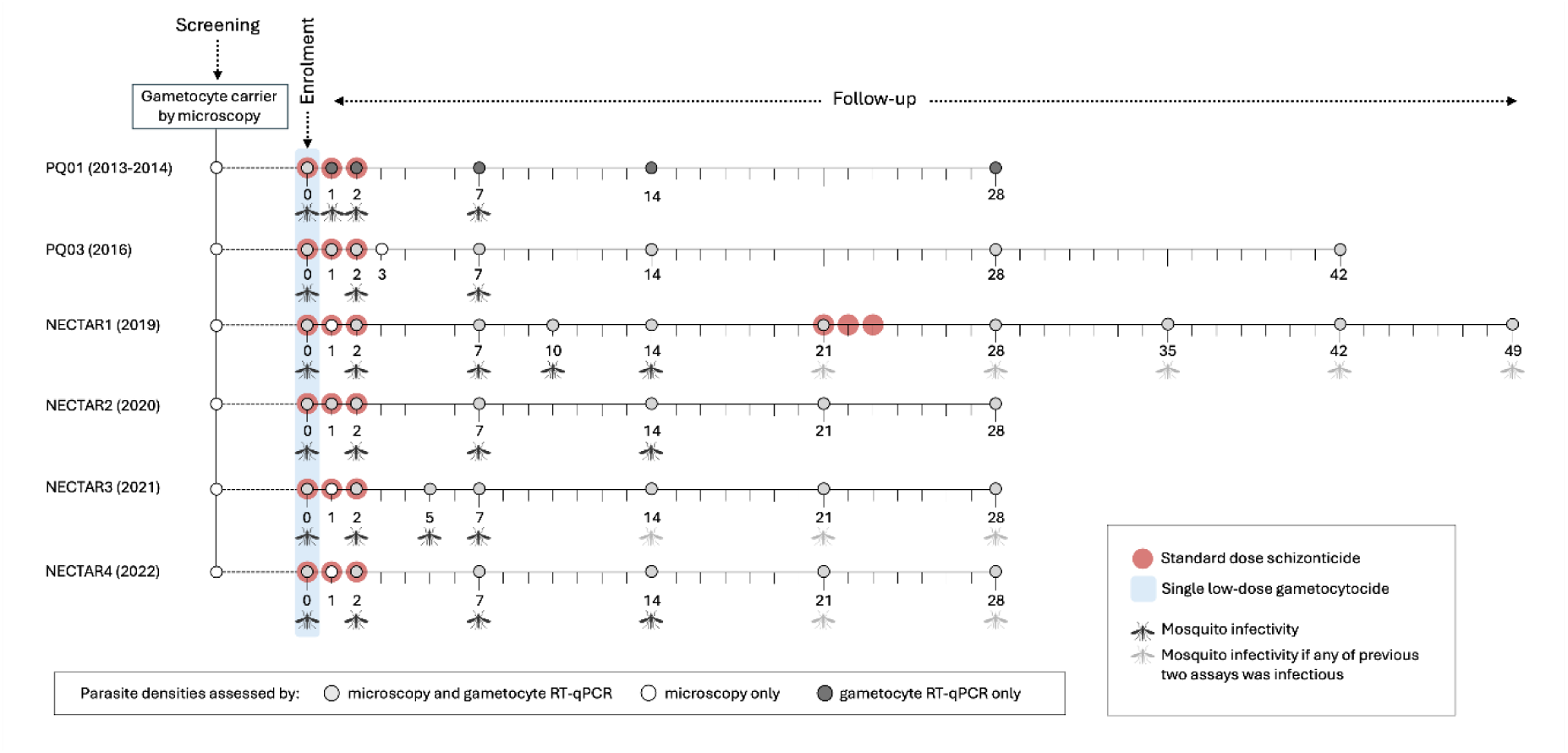
Study design of included trials. All studies assessed infectivity to mosquitoes before (d0), during (d2) and one and two weeks after initiation of treatment (d7 and d14) with additional time-points that differed between studies. Tick marks represent days; circles indicate sampling/screening time points. Circles that are encompassed by larger red circles indicate that a standard dose schizonticide was administered at these study visits, while a single low-dose gametocytocide was administered at the time points with a larger blue circle. Grey coloured circles indicate that parasite densities were assessed by both microscopy (asexual parasite and gametocyte densities) and RT-qPCR (gametocyte densities). At other study visits, parasite densities were determined by microscopy only (white circles) or RT-qPCR only (dark grey circles). Mosquito infectivity assays were conducted at the study visits marked with a black mosquito symbol, while a grey mosquito symbol indicates that mosquito infectivity assays were only conducted at that time point if any of the previous two assays resulted in at least one infected mosquito.

For infectivity assessments in all studies, 75-90 insectary-reared *Anopheles gambiae* females were allowed to feed for 15–20 min on venous blood samples collected in Lithium Heparin tubes and offered in a water-jacketed membrane feeder system, as previously described (27). Fully fed mosquitoes were kept in a temperature and humidity-controlled insectary until day 7 post-feeding, and then dissected in 1% mercurochrome to detect and quantify *P. falciparum* oocysts. Giemsa-stained blood slides were double read by expert research microscopists with asexual stages counted against 200 white blood cells and gametocytes counted against 500 white blood cells. For molecular gametocyte quantification in all studies, venous or finger-prick blood collected in EDTA tubes was aliquoted into L6 buffer (Severn Biotech, Kidderminster, UK) or RNA protect cell reagent (Qiagen, Hilden, Germany) and stored at ≤−70°C until total nucleic acid extraction using a MagNAPure LC automated extractor (Total Nucleic Acid Isolation Kit High Performance; Roche Applied Science, Indianapolis, IN, USA). Male and female gametocytes were quantified in a multiplex reverse-transcriptase quantitative PCR (RT-qPCR) assay targeting Pfs25 or CCP4 transcripts for female gametocytes and PfMGET for male gametocyte quantification, as specified in appendix 2. Molecular gametocyte quantification was repeated for the PQ03 study, after the original density estimates of this study (but not of other studies) showed marked disagreement with their respective microscopy density measurements (supplementary figure 1, appendix 1, p 7). Samples were classified as negative for a specific gametocyte sex if the RT-qPCR quantified gametocyte density was less than 0.01 gametocytes per μL (equivalent to one gametocyte per 100 μL of the blood sample).

### Statistical analysis

Asexual parasite and microscopy gametocyte density distributions at baseline were presented per study by violin plots. Six individuals from the PQ01 study that lacked baseline infectivity measures or microscopy gametocyte densities were excluded from analysis. The association between gametocyte density (gametocytes / µL) on a log10 scale and the proportion of infected mosquitoes was determined by a mixed-effects logistic regression with a random effect for study. The association between (log10) gametocyte density estimates by microscopy and molecular methods was determined by a linear model with an interaction by study.

To quantify reductions in key output parameters (gametocyte prevalence and densities, the proportion of infected mosquitos, mean oocyst density) within treatments over time (days 2, 7 and 14) and between treatments at each time point, mixed-effects generalized linear models were fit with individual-specific random effects and fixed effects for time points, treatment groups and their interaction. A Poisson family with log-link was used for gametocyte prevalence (since a binomial family with log-link failed to converge), a Gamma family with log-link was used for gametocyte densities assessed by microscopy and RT-qPCR, a binomial family with log-link was used for proportion of infected mosquitos, while a negative binomial family with log-link was used for oocyst density.

The models were of the form:

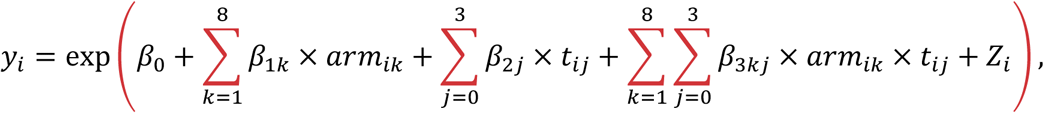

where the study arms were DHA-PPQ, SP-AQ, PY-AS, AS-AQ, AL, Non-ACT-PQ, ACT-PQ and ACT-TQ, and the time points were *t*_0_=day 0, *t*_1_=day 2, *t*_2_=day 7 and *t*_3_=day 14, the *β*′s are the regression parameters to be estimated, *Z_i_* are the individual specific random effects, and *y_i_* is one of the outcomes of interest: gametocyte prevalence or densities, the proportion of infected mosquitos, oocyst density. The beta (*β*) coefficients estimated from the model were used to calculate the relative reductions at each time point for each treatment arm.

Kaplan-Meier survival curves were generated with the *survfit* function from the R-based *survival* package to display the cumulative probability of remaining uncleared of gametocytes detected by microscopy, gametocytes detected by RT-qPCR and any infectivity to mosquitoes. The Cox proportional hazards model was used to estimate hazard ratios and to draw comparisons between treatment arms for each of the clearance events. Statistical analyses and visualisations were conducted in R (version 4.3.2).

The original studies were registered with ClinicalTrials.gov (NCT01743820, NCT02831023, NCT04049916, NCT04609098, NCT05081089 and NCT05550909).

## RESULTS

A total of 422 study participants (42-100 participants per study) and 15 different treatment regimens or dosing schedules were evaluated across the six included trials (20-79 participants per regimen/dosing schedule, Table 1, Appendix 2). Parasite density was broadly similar between studies that recruited asymptomatic parasite carriers (2016–2022) but higher in the PQ01 study conducted in 2013-2014 where clinical malaria patients were eligible (Figure 2A). Although participants were recruited based on microscopically detected gametocyte carriage, at the time of enrolment (up to 24 hours after the initial screening), microscopy gametocyte prevalence was 99.5% (420/422) with a median gametocyte density of 55 (IQR 32-112) gametocytes/µL among gametocyte carriers (Figure 2B). Gametocyte density by microscopy was highly skewed with a range of 15.5-2720 gametocytes/µL (supplementary table 2, appendix 1, p 8). Prior to treatment, 68.8% (290/422) of individuals were infectious to mosquitoes (Figure 2C). The proportion of mosquitoes each participant infected at this time point ranged from 0.75% to 94.3% and was positively associated with microscopically determined gametocyte density (Figure 2D). In the four studies where oocyst numbers were recorded (NECTAR studies 1-4), the proportion of mosquitoes infected was strongly positively associated with mosquito infection burden (oocyst density) (supplementary figure 2, appendix 1, p 9).

**Figure 2.**
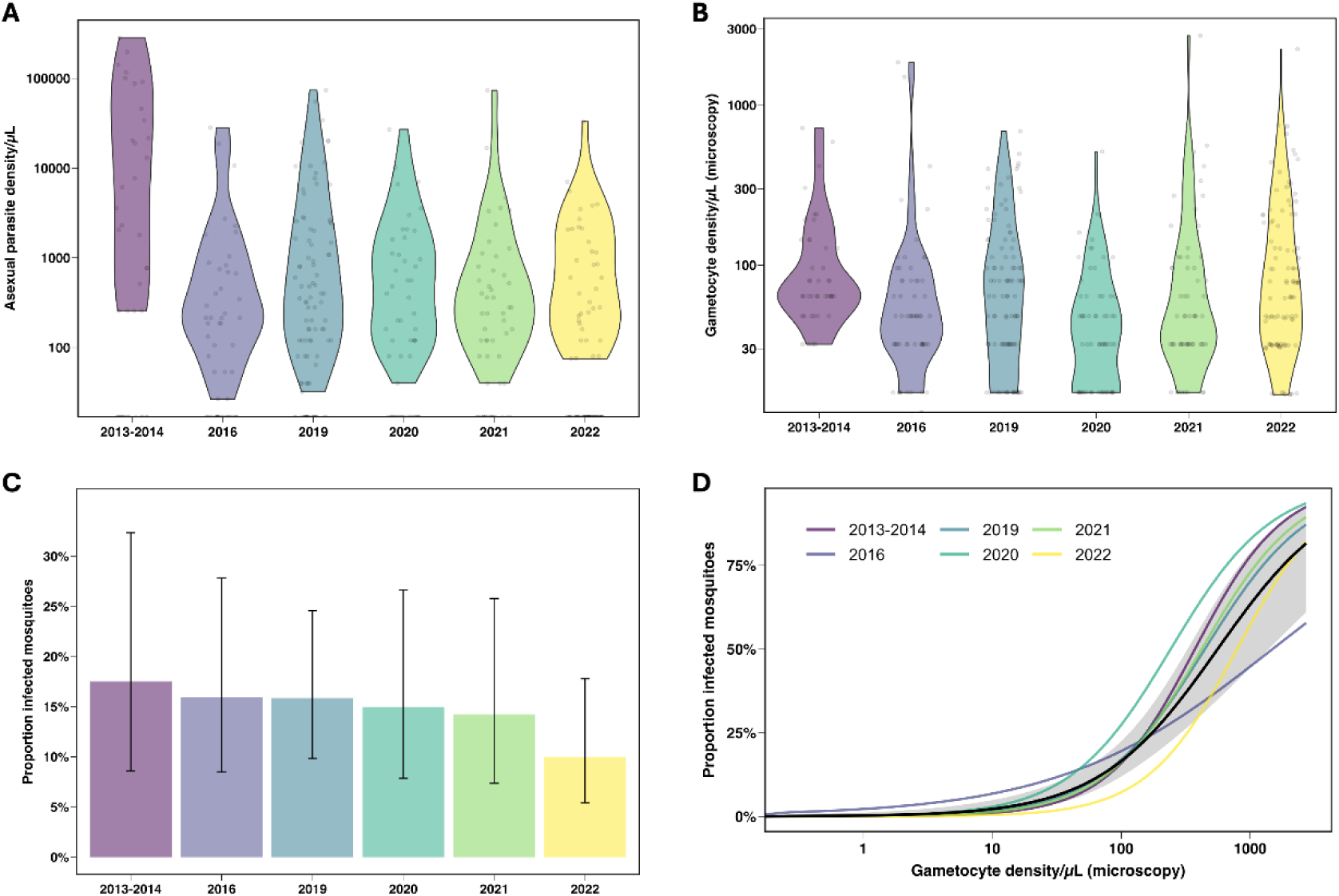
Baseline study characteristics. **A.** Violin plot of microscopical asexual parasite density (parasites/µL) distribution per study. **B.** Violin plot of microscopical gametocyte density (gametocytes/µL) distribution per study. **C**. The mean proportion of mosquitoes that became infected after feeding on venous blood collected at enrolment, prior to treatment, per study. Vertical bars represent 95% CIs estimated from a logistic regression model. **D**. Results from a logistic regression between microscopically determined gametocyte densities (gametocytes / µL) on a log10 scale and the proportion of infected mosquitoes over the different study years shown by the different colours. Mosquito feeding assays in this analysis were conducted before treatment was initiated. The black line indicates the overall trend averaged across all years with the shaded area showing the 95% confidence interval for this overall fit. Visualisations represent a total of 422 observations, from 42 (2013-2014), 60 (2016), 100 (2019), 60 (2020), 60 (2021) and 100 (2022) study participants. The median number of dissected mosquitoes per study participant (panel C) was 71.8 (IQR 65.6-77) for 2013-2014, 79 (IQR 72-84) for 2016, 64 (IQR 57-70) for 2019, 60 (IQR 51.8-66.5) for 2020, 62 (IQR 53.8-64.2) for 2021 and 61 (IQR 55-66) for 2022.

Acknowledging that densities below the microscopic threshold for detection (16 gametocytes/ µL) can result in mosquito infections, gametocyte density was also quantified by molecular methods. For all studies, gametocyte prevalence by molecular methods was 100% at enrolment and there was a strong positive association between gametocyte density by microscopy and by molecular methods (Figure 3A). At enrolment, two study participants were positive for gametocytes by molecular methods but not by microscopy; both became gametocyte positive by microscopy one day after baseline.

**Figure 3.**
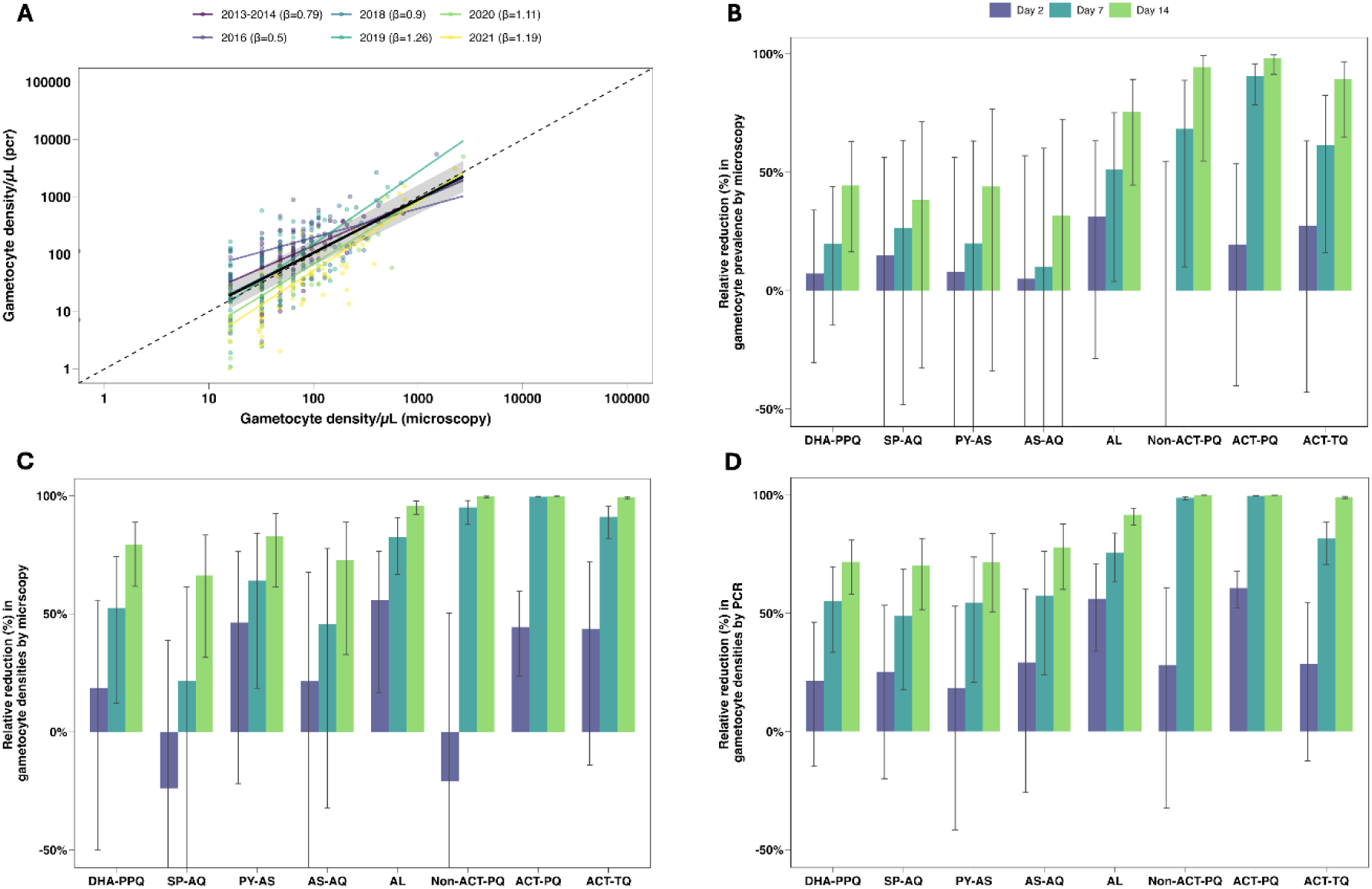
Gametocyte prevalence and densities. **A.** scatter plot of the association between gametocyte density measured by microscopy (x-axis) and RT-qPCR (y-axis) pre-treatment, both on a log10 scale. A linear model with an interaction by year was used to estimate the study specific trend lines (shown in colour with estimated slopes (β) indicated in the legend). A linear mixed effects model was used to estimate the trend line averaged across the study years (β=0.94) shown by the black line, and the corresponding 95% CI, shown by the grey shaded area. The dashed line represents the line of equality where measurements by microscopy and PCR would be equal. **B-D.** Bar charts illustrating the relative reduction compared to baseline in gametocyte prevalence by microscopy (**B**), and gametocyte densities measured by microscopy (**C**) and molecular methods (**D**), by treatment arm over three time points (Day 2 - indigo, Day 7 - turquoise, and Day 14 - green). Vertical bars depict the 95% confidence intervals for these estimates. Due to inflated standard errors the y-axis was cut off below -50. Visualisations represent data from 422 individuals at baseline (79, 40, 25, 20, 60, 20, 138 and 40 individuals from the DHA-PPQ, SP-AQ, PY-AS, AS-AQ, AL, non-ACT-PQ, ACT-PQ and ACT-TQ groups, respectively). At day 2, 369 individuals were included (65, 39, 23, 20, 58, 19, 105 and 40 individuals from the DHA-PPQ, SP-AQ, PY-AS, AS-AQ, AL, Non-ACT-PQ, ACT-PQ and ACT-TQ groups, respectively). Data from 357 individuals at day 7 is shown (57, 38, 24, 20, 57, 19, 104 and 38 individuals from the DHA-PPQ, SP-AQ, PY-AS, AS-AQ, AL, Non-ACT-PQ, ACT-PQ and ACT-TQ groups, respectively). Day 14 includes data from 357 individuals (60, 38, 24, 19, 57, 18, 105 and 36 individuals from the DHA-PPQ, SP-AQ, PY-AS, AS-AQ, AL, Non-ACT-PQ, ACT-PQ and ACT-TQ groups, respectively).

Following treatment, gametocyte prevalence and densities declined in all treatment groups. The extent and speed at which this occurred depended strongly on the treatment provided (Figure 3B-D, supplementary figure 3, appendix 1, p 10). Two days after treatment, relative reductions compared to baseline in microscopically detected gametocyte prevalence were non-significant in all arms. However, at seven days after treatment initiation, this reduction was statistically significant in the AL group (51.12%, 95% CI 3.85% -75.15%, p=0.0381), but not after the other schizonticidal regimens without addition of a gametocytocide (p=0.2258, p=0.3892, p=0.5731 and p=0.7997 after DHA-PPQ, SP-AQ, PY-AS and AS-AQ treatment, respectively). The reduction in gametocyte prevalence was more rapid for treatment groups that included a single low-dose gametocytocide, with reductions of 68.31% (95% CI 10.02% -88.84%, p=0.0309) in the non-ACT-PQ group, 90.52% (95% CI 78.52% -95.81%, p<0.0001) in the ACT-PQ group and 61.53% (95% CI 16.07% -82.37%, p=0.0164) in the ACT-TQ group, at day 7 compared to baseline. Relative reductions in gametocyte prevalence assessed by microscopy and molecular methods at days 2, 7 and 14 post-treatment are presented in Figure 3B and supplementary tables 3-8 (appendix 1, pp 11-16). We combined different ACTs with PQ for this analysis; the findings per study arm are given in supplementary figure 4 (appendix 1, p17).

Reductions in gametocyte prevalence were predictably slower than reductions in gametocyte densities, which showed significant relative reductions at day 2 compared to baseline by both microscopy and molecular measures in the AL (p=0.0116 and p<0.0001, respectively) and ACT-PQ (p=0.0003 and p<0.0001, respectively) groups (Figure 3 C-D, supplementary tables 9-14, supplementary figure 5, appendix 1, pp 18-24). Reductions in PCR-determined gametocyte density on day 2 after initiation of treatment were more pronounced for AL compared to DHA-PPQ (p=0.0036), SP-AQ (p=0.0308), PY-AS (p=0.0302) (supplementary table 12, appendix 1, p 21) with similar patterns at day 7 and 14 (supplementary tables 13 and 14, appendix 1, p 22-23). AS-AQ also resulted in a smaller reduction in gametocyte density compared to AL, which was not significant on day 2 (p=0.1085) but became significant at later timepoints (p=0.0617 and p=0.0013 at days 7 and 14). ACT-PQ resulted in the fastest reductions in molecular gametocyte density and this reduction was significantly larger than the best performing ACT, AL, on days 7 and 14 (p<0.0001; supplementary tables 13 and 14, appendix 1, pp 22-23). Non-ACT-PQ and ACT-TQ regimens showed reductions of nearly 30% in gametocyte density by PCR on day 2, which was significantly less than in the ACT-PQ group (p=0.0400 and p=0.0043, respectively). Across all post-treatment timepoints, the proportion of gametocyte infections that were submicroscopic was 28.63% (777/2714).

A valuable feature of the studies included was the availability of mosquito infection data before and after initiation of treatment. In all groups, 58.3% to 95% of participants infected mosquitoes before treatment (supplementary table 2, appendix 1, p 8), with 99.7% (289/290) of infectious individuals having gametocytes detected by microscopy at this timepoint. Following treatment, as gametocyte densities declined, the contribution of submicroscopic infections to transmission increased (Figure 4A). At day 2, 8.33% (12/144) of all infectious individuals had submicroscopic gametocyte infections. At days 7 and 14, these percentages were 6.59% (6/91) and 3.70% (1/27), respectively. The experiments in which mosquitoes became infected after feeding on samples with submicroscopic gametocyte densities typically resulted in low proportions of infected mosquitoes.

**Figure 4.**
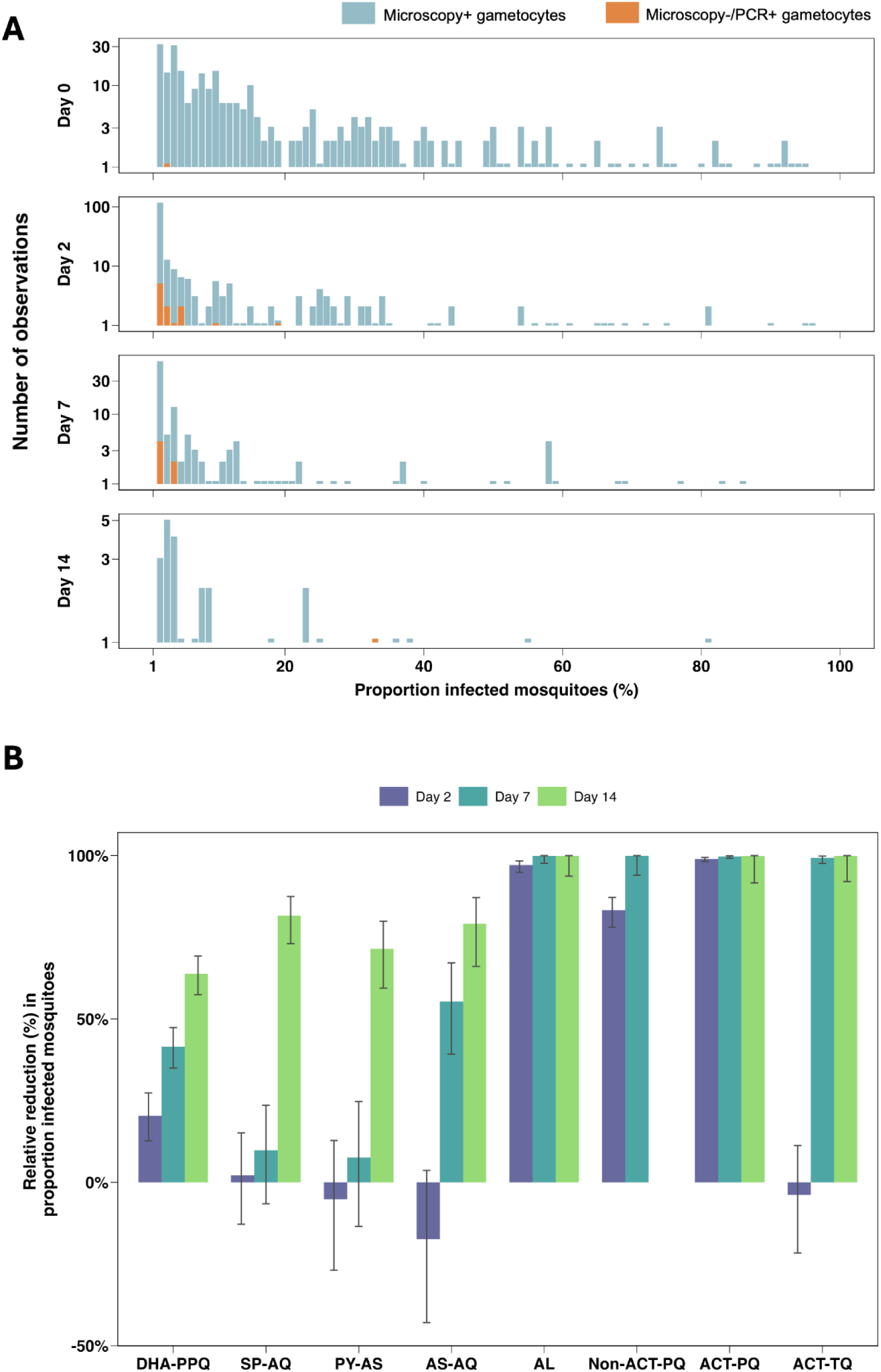
Infectivity of submicroscopic gametocyte infections and the relative reduction in the proportion of infected mosquitoes compared to baseline per treatment category. **A**. Stacked bar chart representing the number of observations for each proportion of infected mosquitoes (rounded to the nearest integer) at baseline and at days 2,7 and 14 post-treatment initiation. For each day of follow-up, individual study participants contribute a single observation. Bars are coloured by the presence of gametocytes by microscopy (blue) or by PCR only (orange). Baseline visualisations represent a total of 422 study participants; per study 42 (2013-2014), 60 (2016), 100 (2019), 60 (2020), 60 (2021) and 100 (2022) participants were enrolled and presented here. At day 2, data from 375 individuals are presented (60, 99, 60, 58, 98 participants from the 2016, 2019, 2020, 2021 and 2022 studies, respectively). At day 7 post-treatment initiation, 367 participants are presented (56, 98, 59, 58 and 97 participants from the 2016, 2019, 2020, 2021 and 2022 studies, respectively). Finally, at day 14, data from 218 individuals are presented (47, 56, 17 and 96 participants from the 2019, 2020, 2021 and 2022 studies, respectively). **B.** Bar chart illustrating the relative reduction compared to baseline in the proportion of infected mosquitoes by treatment arm over three time points (Day 2 - indigo, Day 7 - turquoise, and Day 14 - green). Vertical bars depict the 95% confidence intervals for these estimates. Visualisations represent data from 416 individuals at baseline (79, 40, 25, 20, 60, 20, 139 and 40 individuals from the DHA-PPQ, SP-AQ, PY-AS, AS-AQ, AL, non-ACT-PQ, ACT-PQ and ACT-TQ groups, respectively). At day 2, 416 individuals were included (78, 40, 25, 20, 58, 20, 135 and 40 individuals from the DHA-PPQ, SP-AQ, PY-AS, AS-AQ, AL, Non-ACT-PQ, ACT-PQ and ACT-TQ groups, respectively). Data from 409 individuals at day 7 is shown (76, 39, 25, 20, 57, 19, 134 and 39 individuals from the DHA-PPQ, SP-AQ, PY-AS, AS-AQ, AL, Non-ACT-PQ, ACT-PQ and ACT-TQ groups, respectively). Day 14 includes data from 218 individuals (43, 15, 25, 19, 39, 40 and 37 individuals from the DHA-PPQ, SP-AQ, PY-AS, AS-AQ, AL, ACT-PQ and ACT-TQ groups, respectively).

Transmission blocking effects per treatment group were quantified as averages of individual level effects; that is, within-person reductions in the proportion of mosquitoes infected at post-treatment timepoints compared to baseline (Figure 4B, supplementary figures 6 and 7, appendix 1, pp 28-29). All treatment groups showed a statistically significant relative reduction in mosquito infection rates by day 14 (Figure 4B, supplementary tables 15-17, appendix 1, pp 25-27). At earlier time-points, we observed a modest but statistically significant reduction in mosquito infection prevalence on day 2 for DHA-PPQ (20.35%, 95% CI 12.70-27.33, p<0.0001) and for non-ACT-PQ (83.26%, 95% CI 78.11-87.21, p<0.0001) whilst >90% transmission reduction was observed for AL and for ACTs with SLD PQ. For AS-AQ and ACT with SLD TQ, a statistically significant reduction in mosquito infection rates was only observed at day 7, while for SP-AQ and PY-AS, this was only observed by day 14. For these treatment groups, relative differences in infection rates at day 2 ranged from reductions of 2.17% in the SP-AQ arm (95% CI -12.80% -15.14%) to increases in infectivity in the AS-AQ arm (‘reduction’ of -17.32%, 95% CI -42.92-3.69); neither of these findings were statistically significant. Taken together, AL performed significantly better at reducing mosquito infection rate within 48 hours of treatment, compared to the other ACT regimens DHA-PPQ, PY-AS and AS-AQ at day 2 (p<0.0001 compared to AL) (Figure 5A). The differences in reductions in infectivity became overall smaller at day 7 after treatment (Figure 5B), however, SP-AQ and PY-AS still only showed a <10% reduction, while the reduction in mosquito infection was increased in the DHA-PPQ and AS-AQ groups to 41.51% (95% CI 34.98% - 47.37%) and 55.32% (95% CI 39.21% - 67.16%), respectively

**Figure 5.**
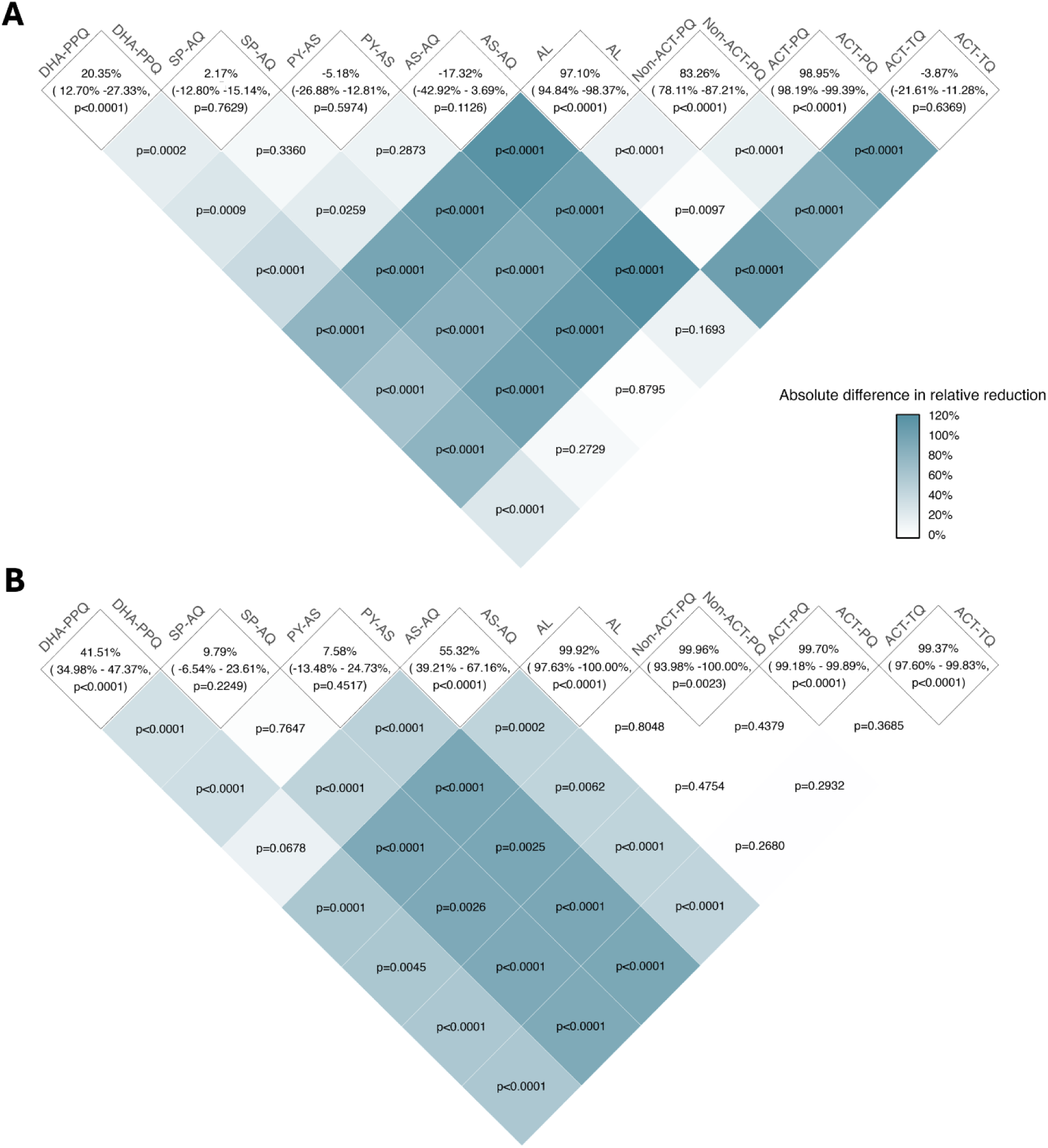
Between-group comparison of the reduction in proportion infected mosquitoes at days 2 and 7 post-treatment, compared to baseline. Heatmaps representing the percentage reduction in the proportion infection mosquitoes per treatment category at day 2 (**A**) and day 7 (**B**) compared to baseline in the top cells, with 95% confidence intervals and p-value comparing to baseline. Other cells in the heatmap represent the p-values comparing treatment groups. Heatmap cells are coloured by the absolute difference between treatment groups in the relative reduction in proportion infected mosquitoes at days 2 and 7 post-treatment, compared to baseline. For example, DHA-PPQ achieves 20.35% reduction in the proportion of infected mosquitoes by day 2 compared to 97.10% for AL; the difference between these arms (76.75% lower reduction for DHA-PPQ) is statistically significant (p<0.0001).

Over all treatment groups, 27 individuals became infectious to mosquitoes after treatment while initially not infecting mosquitoes; 13.92% (11/79) of participants in DHA-PPQ groups, 12.5% (5/40) in SP-AQ groups, 16% (4/25) in the PY-AS group, 10% (2/20) in the AS-AQ group, 1.67% (1/60) in AL groups, 0.72% (1/138) in the ACT plus primaquine groups and 7.5% (3/40) in ACT plus tafenoquine groups. The probability of infecting at least one mosquito was reduced predictably slower than the mosquito infection rate, with only the groups with PQ reaching >90% reduction in this probability at day 2, followed by AL that achieved >80% reduction at day 7 and ACT-TQ that achieved this at day 14 (supplementary Figure 8, supplementary Tables 18-20, appendix 1 pp 30-33). Relative reductions in oocyst density in dissected mosquitoes preceded the reduction in mosquito infection rates in certain treatment groups, such as DHA-PPQ, SP-AQ, PY-AS and AS-AQ, where a larger reduction in oocyst density was found prior to observing the same level of reduction in mosquito infection rate (supplementary Figure 9, supplementary tables 21-23, appendix 1 pp 34-37).

The clearance of infectivity (i.e. no longer infecting any mosquitoes) preceded the clearance of gametocytes assessed by both microscopy and RT-qPCR (Figure 6). The treatment groups with SLD PQ exhibited the fastest infectivity clearance, followed by those treated with AL (hazard ratios 0.62 (p<0.0001) and 0.66 (p=0.0006) compared to non-ACT-PQ and ACT-PQ, respectively). Infectivity was annulled by day 7 in the AL, ACT-PQ and non-ACT-PQ groups, and by day 14 in the ACT-TQ group, while this effect was slower in the SP-AQ (day 28), DHA-PPQ (day 35) and PY-AS (day 35) groups. Not all infectivity was cleared by the end of follow up (day 28) in the AS-AQ group (4.55 % of individuals still infectious, 95% CI 0.67-30.85). Hazard ratios and between-arm comparisons for the clearance of infectivity, adjusted by baseline PCR gametocyte densities, can be found in supplementary table 24 (appendix 1 p 38). ACT-PQ was significantly faster than all other regimens at gametocyte clearance (p<0.0001 in all comparisons; supplementary tables 25-26, appendix 1 pp 39-40). Gametocytes detectable by microscopy persisted after day 28 in the DHA-PPQ, SP-AQ, PY-AS, AL and non-ACT-PQ groups, while gametocytes were microscopically undetectable by day 14 in the ACT-PQ group and by day 21 in the ACT-TQ group. Gametocytes measured by RT-qPCR (limit of detection of 0.01 gametocytes/µL) persisted in a subset of individuals in all treatment groups until the end of follow-up.

**Figure 6.**
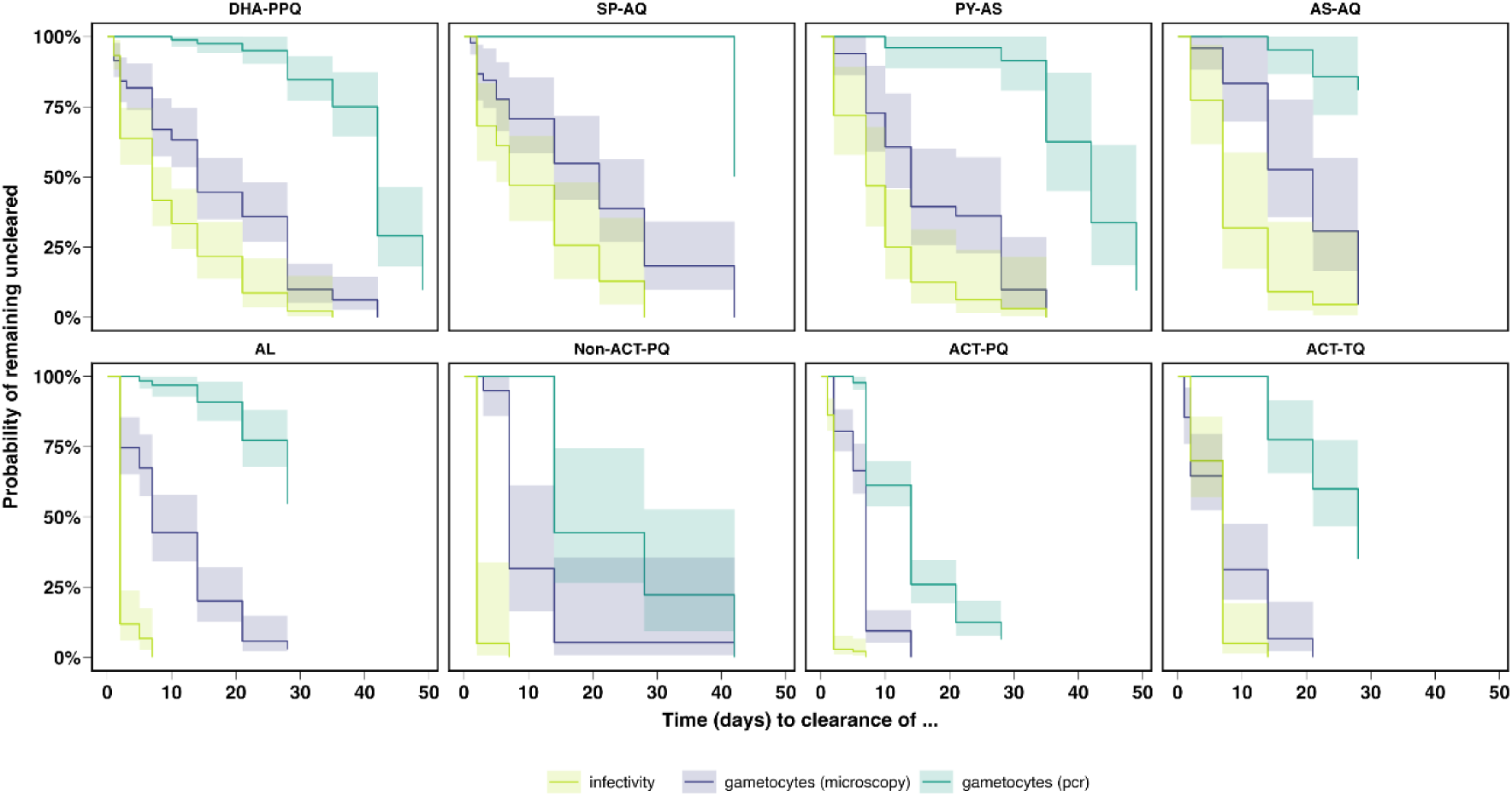
Time to clearance of infectivity and gametocytes per treatment category. Kaplan-Meier survival curves showing the cumulative probability of remaining uncleared of gametocytes detected by microscopy (purple), gametocytes detected by PCR (turquoise) and infectivity to mosquitos (green) over time stratified across different antimalarial treatment categories (DHA-PPQ, SP-AQ, PY-AS, AS-AQ, AL, Non-ACT-PQ, ACT-PQ, ACT-TQ). Shaded areas indicate 95% confidence intervals for the Kaplan-Meier survival estimates. Survival curves showing infectivity represent data from 417 individuals (79, 40, 25, 20, 58, 20, 135 and 40 individuals from the DHA-PPQ, SP-AQ, PY-AS, AS-AQ, AL, Non-ACT-PQ, ACT-PQ and ACT-TQ groups, respectively). Survival curves visualising gametocytes by microscopy show data from 375 individuals (65, 40, 25, 20, 58, 20, 107 and 40 individuals from the DHA-PPQ, SP-AQ, PY-AS, AS-AQ, AL, Non-ACT-PQ, ACT-PQ and ACT-TQ groups, respectively) and 417 individuals (79, 40, 25, 20, 58, 20, 135 and 40 individuals from the DHA-PPQ, SP-AQ, PY-AS, AS-AQ, AL, Non-ACT-PQ, ACT-PQ and ACT-TQ groups, respectively) for gametocyte assessment by RT-qPCR.

## DISCUSSION

This pooled analysis from individual patient data compares the transmission-blocking activity of different regimens of schizonticidal drugs with or without a single low-dose gametocytocide in *P. falciparum* gametocyte carriers in Mali. We found a marked difference between the different ACTs, with AL causing the largest reduction in mosquito infection rate (97.10%) within 48 hours of treatment initiation. The impact of DHA-PPQ, PY-AS and AS-AQ on transmission were significantly less than AL and led to prolonged gametocyte carriage and infectivity post-treatment. Adding a single low-dose of primaquine (0.25 or 0.5 mg/kg) to any ACT accelerated the clearance of gametocytes and led to a significantly greater reduction in mosquito infection rate within 48-hours of treatment.

The effectiveness and utility of the treatments in the six studies analysed here have been discussed in depth in the original trial reports (21–26). This combined analysis represents the first synthesis of this data and direct comparison of these treatments. A number of treatments were tested in multiple studies (DHA-PPQ, DHA-PPQ plus 0.25 mg/kg PQ, AL and SP-AQ) and our results were highly concordant across years (supplementary figure 7, appendix 1, p 29), evidence of the value of established transmission testing facilities and the appropriateness of a pooled analysis. Prior to these trials, there was sparse evidence of the gametocytocidal and transmission reducing efficacy for many of the included treatments. The available data frequently originated from single studies, used disparate and insensitive methodologies for gametocyte quantification, or did not conduct transmission assays to determine the transmissibility of gametocytes (28–32). This was the rationale for including only studies with highly similar enrolment criteria and assessments, including transmission assays before and after initiation of treatment.

This pooled analysis confirms SP-AQ’s poor ability to clear gametocytes and reduce transmission, with a significantly lower relative reduction in mosquito infection rate at day 7 compared to DHA-PPQ, AS-AQ, and AL. As the only non-artemisinin-based antimalarial recommended for systematic distribution via seasonal malaria chemoprevention (SMC) (33), SP-AQ’s poor performance against gametocytes may reduce the overall community impact of SMC; adding a single low-dose of primaquine to SP-AQ for chemoprevention, would likely improve this.

This pooled analysis provides the first direct comparison of SLD primaquine and SLD tafenoquine with the same partner drug. Our analysis indicates a delayed but highly effective response of SLD tafenoquine compared to SLD primaquine when combined with ACTs, achieving a 99.37% reduction in mosquito infection rate at day 7 from baseline. Although the transmission-blocking efficacy of tafenoquine is dose-dependent, our pooled analysis combined the two highest dose groups (0.83 mg/kg and 1.66 mg/kg) for simplicity, possibly obscuring the effects of the higher dose. Despite these delayed transmission-blocking properties of TQ compared with SLD primaquine in our findings, its long half-life could be a major advantage and could be of relevance to prevent the transmission of drug resistant parasites, which may have an increased gametocyte conversion rate and transmission potential (34,35).

Before ACTs were introduced, several antimalarials including chloroquine and sulfadoxine-pyrimethamine were reported to increase gametocytaemia and/or infectivity after treatment (36,37). Our analysis, almost exclusively examining ACTs, did not find evidence for post-treatment enhancement of infectivity although we did observe that a non-negligible proportion of individuals became infectious after treatment (this proportion being ≥10% of participants in the DHA-PPQ, SP-AQ, PY-AS and AS-AQ groups); this percentage was below 2% in the remaining treatment groups, including AL. The fact that very few individuals become gametocyte positive or infectious after AL suggests that, among ACT regimens, AL may be superior at targeting immature or sequestered gametocytes.

The presence of gametocytes after treatment at densities that normally permit transmission does not make transmission an inevitability, because gametocytes may be sterilized or become non-infectious for other reasons before being cleared (12). In this pooled analysis, this discordance was most pronounced in the AL group and groups with SLD PQ, where post-treatment effects on infectivity preceded gametocyte clearance. In all other treatment groups, reductions in gametocyte density and mosquito infection rate followed more similar patterns. The combination of an ACT with SLD PQ was significantly faster than all other regimens at gametocyte clearance (100% of microscopical gametocytes by day 14), while ACT-TQ was the second fastest regimen to clear gametocytes (100% of microscopical gametocytes by day 21). Infectivity clearance was the fastest in the treatment groups with SLD PQ, followed by AL.

A few limitations in our study warrant consideration. Firstly, we established transmission-blocking activity in highly infectious individuals. This population allows a detailed assessment of transmission-blocking properties; however, post-treatment transmission potential is likely to be smaller in the majority of malaria patients who predominantly have submicroscopic gametocyte densities or may even be free of gametocytes at clinical presentation. A different study would be needed to determine the relative importance of sub-vs microscopic gametocyte carriage for malaria transmission in clinical patients though these would likely require larger sample sizes for mosquito infectivity endpoints. Secondly, our study utilized direct membrane feeding assays rather than direct skin feeding, which may have resulted in reduced infectivity, despite no compelling evidence of gametocytes sequestering in the skin (38). In addition, we measured oocyst prevalence instead of sporozoite prevalence and some infected mosquitoes may not have become infectious (39). Nevertheless, comparisons between groups remain valid, and we observed a strong association between oocyst prevalence and oocyst densities, indicating that a higher proportion of infected mosquitoes reflects a greater transmission potential. Lastly, this pooled analysis set out to assess the ability of antimalarial treatment to prevent the transmission of drug-resistant parasites. Considering that gametocytes in *PfKelch13* mutant infections might preferentially survive artemisinin exposure and infect mosquitoes (35), our findings regarding ACTs may not be generalisable to areas where artemisinin partial resistance is present. In addition, increased gametocytaemia was found to be an early indicator of resistance emergence against previous non-ACT first-line treatments (34). Consequently, while our results indicate that AL is nearly as effective as ACT-PQ in reducing mosquito infection rates, it remains unclear whether this would extend to settings with artemisinin-resistant malaria infections. Our data on SLD gametocytocides in combination with ACTs therefore supports the 2023 advice from the WHO malaria policy and advisory group to expand the focus on reducing parasite transmission with SLD PQ in areas where partial artemisinin resistance has been detected (20). Additionally, we observed high transmission-blocking efficacy of AL when administered as a directly observed therapy (DOT); its efficacy may be lower in real-life settings where treatment adherence may be lower.

In conclusion, utilizing individual patient-level data from six clinical trials conducted at the same study site with highly consistent transmission experiments across trials, we showed pronounced differences in anti-gametocyte and anti-transmission effects between ACTs, with AL showing the most effectiveness in blocking transmission. Additionally, our findings confirm the rapid effects of SLD PQ in clearing and sterilizing gametocytes when used in combination with an ACT, while the addition of an SLD TQ to ACTs has a delayed transmission-blocking effect compared to SLD PQ. Lastly, our analysis confirms considerably higher post-treatment transmission after SP-AQ compared to most ACTs, and adding an SLD PQ to SP-AQ may be beneficial to block malaria transmission in community treatment campaigns.

## Supporting information

Supplemental Table - description of studies

## Abbreviations

ACT: artemisinin-based combination therapy
AL: Artemether-lumefantrine
AL-AQ: Artemether-lumefantrine-amodiaquine
AQ: Amodiaquine
AS-AQ: Artesunate-Amodiaquine
DHA-PPQ: Dihydroartemisinin-Piperaquine
G6PD: glucose-6-phosphate dehydrogenase
PY-AS: Pyronaridine-Artesunate
PQ: Primaquine
SLD: Single low-dose
RT-qPCR: reverse-transcriptase quantitative PCR
SP-AQ: Sulfadoxine-pyrimethamine plus Amodiaquine
TACT: Triple artemisinin-based combination therapy
TQ: Tafenoquine
WHO: World Health Organisation.

## Data Availability Statement

All trial data are available through a novel open-access clinical epidemiology database resource, ClinEpiDB; and code for analysis and the selection of data used for this analysis can be found on https://github.com/leenvh/NECTAR-pooled-analysis.

## Acknowledgements

We would like to thank everyone involved in the original studies, including the local safety monitor, members of the data safety and monitoring board, and all MRTC study staff for their assistance and support. Finally, we are thankful to all study participants and the population of Ouélessébougou, Mali, for their cooperation.

## Author contributions

LNV, JR, AM, CD, AD, WS and TB conceived the study. AM, MJS, MER, LNV, KS, YS, SMN, MD, SM, SK, SS, AY, HD, SFT, RG, JMB, CD, AD, WS and TB conceived and undertook individual studies. KL performed molecular analyses. JR, LNV, AM, RG, JMB, CD, AD, WS and TB analysed the data and interpreted results. LNV, JR, WS and TB wrote the first draft of the manuscript. All authors reviewed and edited the manuscript and read and approved the final manuscript.

