## Supplemental file 1 for "The transmission blocking activity of artemisinin-combination, non-artemisinin, and 8-aminoquinoline antimalarial therapies: a pooled analysis of individual participant data"

**Appendix 1**

### Supplementary Information 1. Antimalarial treatment dosage

*Sulphadoxine-Pyrimethamine plus Amodiaquine*

SP tablets containing 500 mg sulfadoxine and 25 mg pyrimethamine and AQ tablets containing 150 mg amodiaquine were administered according to weight as per manufacturer guidelines shown below:

| **Body weight** | **500/50 mg sulfadoxine/pyrimethamine tablet** | | |
| --- | --- | --- | --- |
|  | **Day 0** | **Day 1** | **Day 2** |
| **11 to 20 kg** | 1x 1 tablet | 1x 1 tablet | 1x 1 tablet |
| **21 to 30 kg** | 1x 1·5 tablets | 1x 1·5 tablets | 1x 1·5 tablets |
| **31 to 45 kg** | 1x 2 tablets | 1x 2 tablets | 1x 2 tablets |
| **> 45 kg** | 1x 3 tablets | 1x 3 tablets | 1x 3 tablets |
| **Body weight** | **150 mg amodiaquine tablet** | | |
|  | **Day 0** | **Day 1** | **Day 2** |
| **15 to 18 kg** | 1x 1·5 tablets | 1x 1 tablet | 1x 1 tablet |
| **19 to 24 kg** | 1x 1·5 tablets | 1x 1·5 tablets | 1x 1·5 tablets |
| **25 to 35 kg** | 1x 2·5 tablets | 1x 2·5 tablets | 1x 2 tablets |
| **36 to 50 kg** | 1x 3 tablets | 1x 3 tablets | 1x 3 tablets |
| **> 50 kg** | 1x 4 tablets | 1x 4 tablets | 1x 3 tablets |

*Artemether-lumefantrine*

AL treatment tablets containing 20/120 mg artemether/lumefantrine or 80/480 mg artemether/lumefantrine were administered according to weight as per manufacturer guidelines shown below:

| **Body weight (kg)** | **20/120 mg artemether/lumefantrine tablet** | | | **80/480 mg artemether/lumefantrine tablet** | | |
| --- | --- | --- | --- | --- | --- | --- |
|  | **Day 0** | **Day 1** | **Day 2** | **Day 0** | **Day 1** | **Day 2** |
| **5 to < 15 kg** | 2x 1 tablet | 2x 1 tablet | 2x 1 tablet | - | - | - |
| **15 to < 25 kg** | 2x 2 tablets | 2x 2 tablets | 2x 2 tablets | - | - | - |
| **25 to < 35 kg** | 2x 3 tablets | 2x 3 tablets | 2x 3 tablets | - | - | - |
| **≥ 35 kg** | 2x 4 tablets | 2x 4 tablets | 2x 4 tablets | 2x 1 tablet | 2x 1 tablet | 2x 1 tablet |

*Dihydroartemisinin-Piperaquine*

Treatment tablets containing 160/320 mg piperaquine with 20/40 mg dihydroartemisinin tablets were administered according to weight as per manufacturer guidelines shown below:

| **Body weight (kg)** | **Total daily dose (mg)**  **(1x/day for 3 days)** | | **Tablet strength and number of tablets per dose** |
| --- | --- | --- | --- |
|  | Piperaquine | DHA |  |
| 5 to <7 | 80 | 10 | ½ x 160mg / 20mg |
| 7 to <13 | 160 | 20 | 1 x 160mg / 20mg |
| 13 to <24 | 320 | 40 | 1 x 320mg / 40mg |
| 24 to <36 | 640 | 80 | 2 x 320mg / 40mg |
| 36 to <75 | 960 | 120 | 3 x 320mg / 40mg |
| 75 to 80 | 1,280 | 160 | 4 x 320mg / 40mg |
| >80 | Not eligible | | |

*Artesunate-Amodiaquine*

Tablets contained 50mg/135 mg or 100mg/270 mg of artesunate/amodiaquine and were administered according to manufacturer guidelines, as shown below:

| **Weight** | **Tablets** | **D0** | **D1** | **D2** |
| --- | --- | --- | --- | --- |
| 9 to < 18 kg | 50 mg AS/135 mg AQ base | 1 tab | 1 tab | 1 tab |
| 18 to < 36 kg | 100 mg AS/270 mg AQ base | 1 tab | 1 tab | 1 tab |
|  | blister pack of 3 tab |  |  |  |
| ≥ 36 kg | 100 mg AS/270 mg AQ base | 2 tab | 2 tab | 2 tab |
|  | blister pack of 6 tab |  |  |  |

*Pyronaridine-Artesunate*

PY-AS granules containing 60 mg pyronaridine-tetraphosphate/20mg artesunate were administered to children <20kg, and PY-AS tablets containing 180 mg pyronaridine-tetraphosphate/60mg artesunate were administered to children and adults >20kg, according to weight as per manufacturer guidelines shown below:

| **Granules (Children <20kg)** | | | |
| --- | --- | --- | --- |
| **Body weight (kg)** | **Total daily dose (mg)**  **(1x/day for 3 days)** | | **Sachet strength and number of tablets per dose** |
|  | Pyronaridine-tetraphosphate | Artesunate |  |
| 5 - <8kg | 60 | 20 | 1 x 60mg/20mg |
| 8 - <15kg | 120 | 40 | 2 x 60mg/20mg |
| 15 - <20kg | 180 | 60 | 3 x 60mg/20mg |

| **Tablets (Children and adults >20kg)** | | | |
| --- | --- | --- | --- |
| **Body weight (kg)** | **Total daily dose (mg)**  **(1x/day for 3 days)** | | **Tablet strength and number of tablets per dose** |
|  | Pyronaridine-tetraphosphate | Artesunate |  |
| 20 - <24kg | 180 | 60 | 1 x 60mg/20mg |
| 24 - <45kg | 360 | 120 | 2 x 60mg/20mg |
| 45-<65kg | 540 | 180 | 3 x 60mg/20mg |
| >65kg | 720 | 240 | 4 x 180mg/60mg |

*Primaquine*

Primaquine tablets were dissolved to a 1 mg/mL solution in distilled water and administered orally to the nearest mL, according to bodyweight at 0·25 mg/kg. Primaquine was administered as a single dose immediately after the first dose of ACT.

*Tafenoquine*

100mg Tafenoquine tablets were available for this study, and were prepared into a 1mg/mL solution in water for weight-based dosing in 5 kg bands as follows:

**1.66 mg/kg Tafenoquine**

| **Weight min** | **Weight max** | **TQ 1mg/mL total (mL)** | **Water (mL)** | **Masking solution (mL)** |
| --- | --- | --- | --- | --- |
| 30 | 35 | 54.0 | 136.1 | 10 |
| 35.01 | 40 | 62.3 | 127.7 | 10 |
| 40.01 | 45 | 70.6 | 119.4 | 10 |
| 45.01 | 50 | 78.9 | 111.1 | 10 |
| 50.01 | 55 | 87.2 | 102.8 | 10 |
| 55.01 | 60 | 95.5 | 94.5 | 10 |
| 60.01 | 65 | 103.8 | 86.2 | 10 |
| 65.01 | 70 | 112.1 | 77.9 | 10 |
| 70.01 | 75 | 120.4 | 69.6 | 10 |
| 75.01 | 80 | 128.7 | 61.3 | 10 |

**0.83 mg/kg Tafenoquine**

| **Weight min** | **Weight max** | **TQ 1mg/mL total (mL)** | **Water (mL)** | **Masking solution (mL)** |
| --- | --- | --- | --- | --- |
| 30 | 35 | 27.0 | 163.0 | 10 |
| 35.01 | 40 | 31.1 | 158.9 | 10 |
| 40.01 | 45 | 35.3 | 154.7 | 10 |
| 45.01 | 50 | 39.4 | 150.6 | 10 |
| 50.01 | 55 | 43.6 | 146.4 | 10 |
| 55.01 | 60 | 47.7 | 142.3 | 10 |
| 60.01 | 65 | 51.9 | 138.1 | 10 |
| 65.01 | 70 | 56.0 | 134.0 | 10 |
| 70.01 | 75 | 60.2 | 129.8 | 10 |
| 75.01 | 80 | 64.3 | 125.7 | 10 |

### Supplementary Table 1. Antimalarial treatment suppliers

| **Study** | **Study drug** | **Supplier** |
| --- | --- | --- |
| **PQ01** | Primaquine | Sanofi, Laval, QC, Canada |
|  | Dihydroartemisinin-piperaquine (Eurartesim) | Sigma-Tau, Pomezia, Italy |
| **PQ03** | Primaquine | Sanofi, Laval, QC, Canada |
|  | Sulfadoxine-pyrimethamine (Fansidar) | Guilin Pharmaceutical, Shanghai, China |
|  | Amodiaquine | Guilin Pharmaceutical, Shanghai, China |
|  | Dihydroartemisinin-piperaquine (Eurartesim) | Sigma-Tau, Pomezia, Italy |
| **NECTAR1** | Pyronaridine-artesunate (Pyramax) | Shin Poong Pharmaceutical, Seoul, South Korea |
|  | Primaquine | ACE Pharmaceuticals, Zeewolde, the Netherlands |
|  | Dihydroartemisinin-piperaquine (Eurartesim) | Sigma Tau, Gaithersburg, MD, USA |
| **NECTAR2** | Dihydroartemisinin-piperaquine (Eurartesim) | Sigma Tau, Gaithersburg, MD, USA |
|  | Tafenoquine | 60° Pharmaceuticals Ltd, USA |
| **NECTAR3** | Artemether-lumefantrine (Coartem) | Novartis, Basel, Switzerland |
|  | Primaquine | ACE Pharmaceuticals, Zeewolde, the Netherlands |
|  | Sulfadoxine-pyrimethamine plus amodiaquine | Guilin Pharmaceutical, Shanghai, China |
|  | Tafenoquine | 60° Pharmaceuticals Ltd, USA |
| **NECTAR4** | Artemether-lumefantrine | Guilin Pharmaceutical, Shanghai, China |
|  | Amodiaquine | Guilin Pharmaceutical, Shanghai, China |
|  | Primaquine | ACE Pharmaceuticals, Zeewolde, The Netherlands |
|  | Artesunate-amodiaquine | Guilin Pharmaceutical, Shanghai, China |

### Supplementary Figure 1. Original PQ03 molecular gametocyte densities

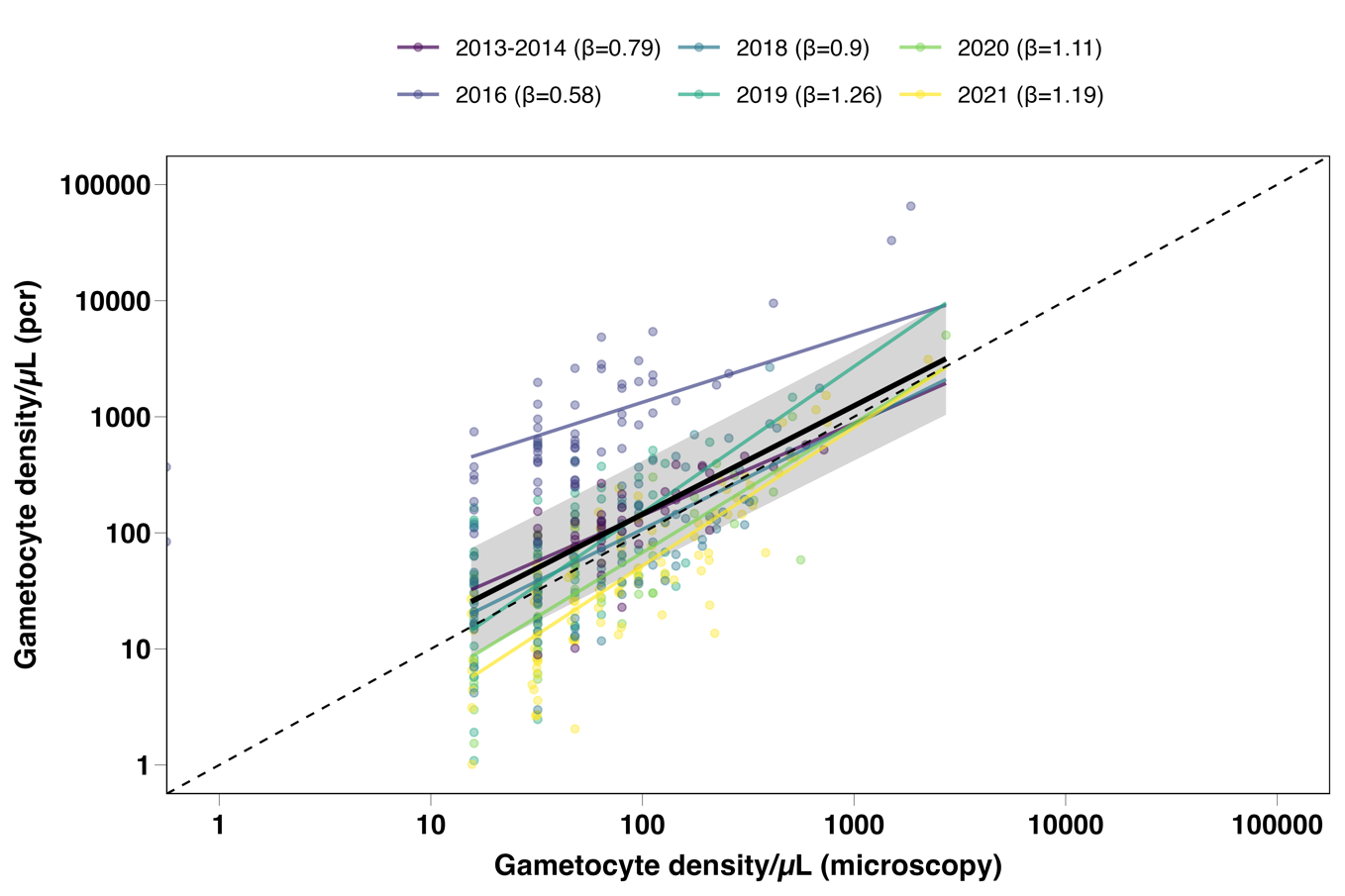

Scatter plot of the association between gametocyte density measured by microscopy (x-axis) and RT-qPCR (y-axis) pre-treatment, both on a log10 scale. The original PQ03 (2016) RT-qPCR data is presented here, which showed that the association with microscopy gametocyte density for this study was an outlier compared to the other studies. Following this, the molecular gametocyte quantification was repeated for this study.

A linear model with an interaction by year was used to estimate the study specific trend lines (shown in colour with estimated slopes (β) indicated in the legend). A linear mixed effects model was used to estimate the trend line averaged across the study years (β=0.94) shown by the black line, and the corresponding 95% CI, shown by the grey shaded area. The dashed line represents the line of equality where measurements by microscopy and PCR would be equal.

### Supplementary Table 2. Baseline descriptives per treatment group

| **Treatment** | **N (%)** | **Males** | **Age** | **Temperature** | **Total parasites/uL** | **Gametocytes/uL (microscopy)** | **Gametocytes/uL (pcr)** | **Female gametocytes/uL (pcr)** | **Proportion infected** | **Gametocyte prevalence** | **Asexual parasite prevalence** | **Proportion participants infectious** |
| --- | --- | --- | --- | --- | --- | --- | --- | --- | --- | --- | --- | --- |
| **DHA-PPQ** | 79 (18.8%) | 59 (74.7%) | 16.5 (10.3) | 36.5 (0.4) | 224.0 (76.0 - 840.0) | 48.0 (16.0 - 80.0) | 80.3 (35.6 - 128.4) | 60.0 (20.8 - 115.7) | 3.0 (0.0 - 14.7) | 77 (97.5%) | 53 (67.1%) | 50 (63.3%) |
| **SP-AQ** | 40 (9.5%) | 29 (72.5%) | 17.2 (11.4) | 36.6 (0.5) | 288.0 (106.0 - 868.0) | 48.0 (32.0 - 112.0) | 65.2 (30.3 - 183.3) | 44.5 (15.1 - 129.2) | 5.2 (0.0 - 10.4) | 40 (100.0%) | 26 (65.0%) | 28 (70.0%) |
| **PY-AS** | 25 (5.9%) | 16 (64.0%) | 13.0 (7.7) | 36.5 (0.6) | 216.0 (64.0 - 704.0) | 64.0 (32.0 - 96.0) | 83.2 (35.9 - 121.7) | 46.0 (25.6 - 74.5) | 3.0 (0.0 - 13.7) | 25 (100.0%) | 13 (52.0%) | 17 (68.0%) |
| **AS-AQ** | 20 (4.7%) | 9 (45.0%) | 14.4 (6.3) | 36.4 (0.3) | 386.3 (162.0 - 1498.4) | 79.0 (41.5 - 232.1) | 52.5 (34.2 - 107.3) | 28.1 (11.3 - 38.9) | 3.9 (1.6 - 17.9) | 20 (100.0%) | 8 (40.0%) | 17 (85.0%) |
| **AL** | 60 (14.2%) | 27 (45.0%) | 18.9 (11.1) | 36.5 (0.4) | 154.5 (48.0 - 411.0) | 48.0 (31.7 - 124.8) | 32.2 (12.8 - 95.8) | 15.6 (6.6 - 38.6) | 3.1 (0.0 - 11.7) | 60 (100.0%) | 29 (48.3%) | 35 (58.3%) |
| **Non-ACT-PQ** | 20 (4.7%) | 20 (100.0%) | 10.3 (3.6) | 36.8 (0.4) | 112.0 (80.0 - 233.3) | 72.0 (32.0 - 96.0) | 198.8 (63.3 - 435.7) | 171.0 (55.1 - 332.6) | 21.6 (4.3 - 33.1) | 20 (100.0%) | 12 (60.0%) | 19 (95.0%) |
| **ACT-PQ** | 138 (32.7%) | 81 (58.7%) | 15.0 (10.0) | 36.4 (0.4) | 328.0 (96.0 - 1473.9) | 70.5 (32.0 - 144.0) | 66.3 (24.8 - 165.6) | 35.5 (12.7 - 109.3) | 5.2 (0.0 - 16.3) | 138 (100.0%) | 83 (60.1%) | 96 (69.6%) |
| **ACT-TQ** | 40 (9.5%) | 28 (70.0%) | 20.2 (10.9) | 36.5 (0.4) | 156.0 (64.0 - 1092.0) | 48.0 (32.0 - 64.0) | 45.2 (19.1 - 204.2) | 22.3 (11.0 - 88.5) | 5.2 (0.0 - 18.8) | 40 (100.0%) | 23 (57.5%) | 28 (70.0%) |

### Supplementary Figure 2. Relation between oocyst density and prevalence

### Supplementary Figure 3. Gametocyte prevalence and density during follow up per treatment category

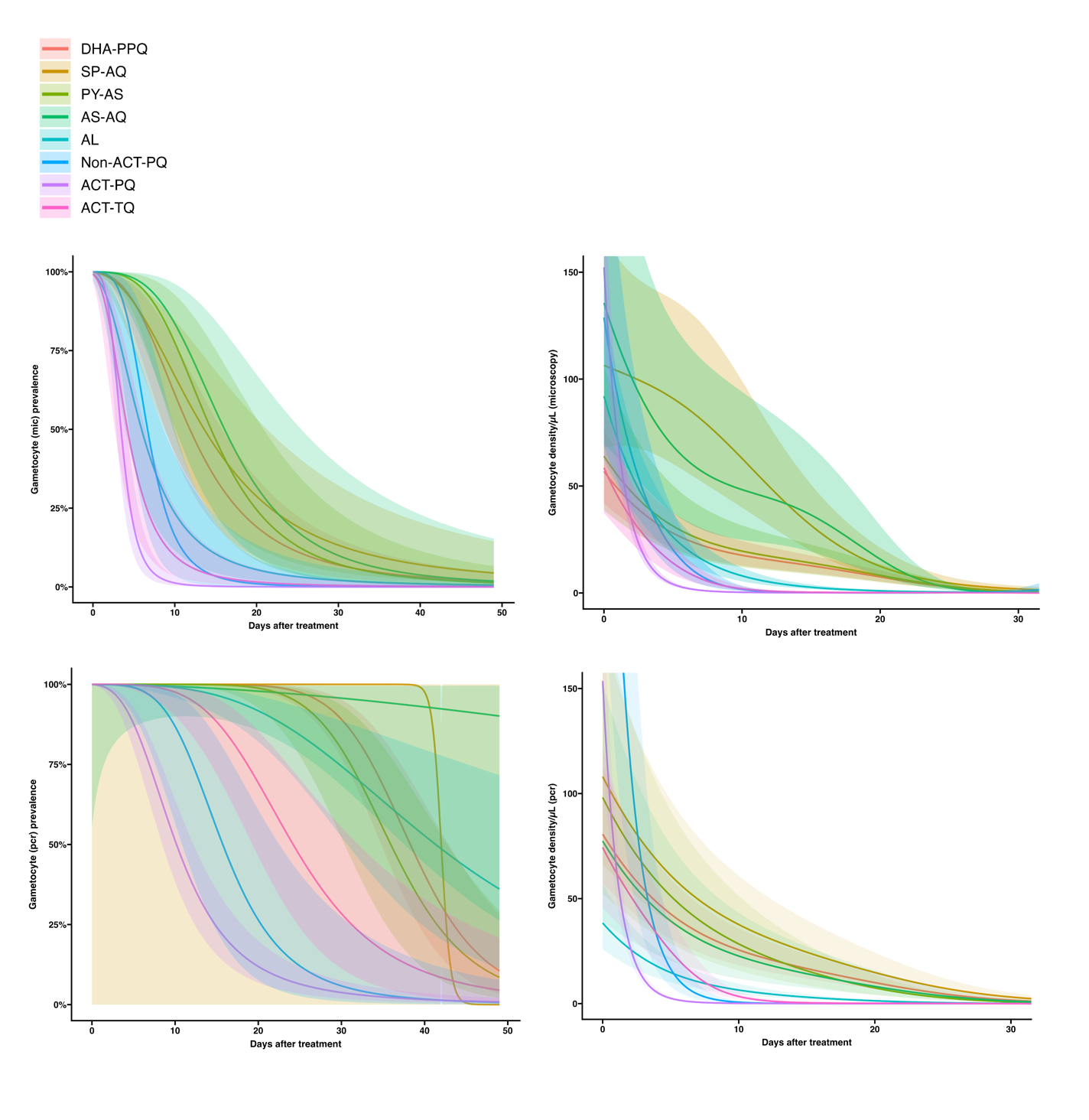

### Supplementary Table 3. Relative reduction in gametocyte prevalence by microscopy at day 2 compared to baseline

| reference | DHA-PPQ | SP-AQ | PY-AS | AS-AQ | AL | Non-ACT-PQ | ACT-PQ | ACT-TQ |
| --- | --- | --- | --- | --- | --- | --- | --- | --- |
| DHA-PPQ | 7.21% (-30.47% -34.01%, p=0.6668) | p=0.7632 | p=0.9799 | p=0.9484 | p=0.2631 | p=0.8356 | p=0.5300 | p=0.4102 |
| SP-AQ | p=0.7632 | 15.00% (-65.17% -56.26%, p=0.6316) | p=0.8312 | p=0.7790 | p=0.4931 | p=0.6792 | p=0.8491 | p=0.6374 |
| PY-AS | p=0.9799 | p=0.8312 | 8.00% (-93.53% -56.27%, p=0.8261) | p=0.9407 | p=0.4098 | p=0.8457 | p=0.6830 | p=0.5287 |
| AS-AQ | p=0.9484 | p=0.7790 | p=0.9407 | 5.00% (-109.65% -56.95%, p=0.8989) | p=0.3942 | p=0.9093 | p=0.6406 | p=0.5020 |
| AL | p=0.2631 | p=0.4931 | p=0.4098 | p=0.3942 | 31.27% (-28.59% -63.26%, p=0.2407) | p=0.3191 | p=0.5131 | p=0.8667 |
| Non-ACT-PQ | p=0.8356 | p=0.6792 | p=0.8457 | p=0.9093 | p=0.3191 | 0.00% (-119.28% -54.40%, p>0.9999) | p=0.5348 | p=0.4207 |
| ACT-PQ | p=0.5300 | p=0.8491 | p=0.6830 | p=0.6406 | p=0.5131 | p=0.5348 | 19.27% (-40.22% -53.52%, p=0.4474) | p=0.7008 |
| ACT-TQ | p=0.4102 | p=0.6374 | p=0.5287 | p=0.5020 | p=0.8667 | p=0.4207 | p=0.7008 | 27.50% (-42.94% -63.23%, p=0.3532) |

Diagonal cells indicate percentage relative reduction at day 2 compared to baseline (95% CI) per treatment category, with corresponding p-value compared to baseline. Other cells in the table contain p-values of between-arm comparisons of the relative reductions.

### Supplementary Table 4. Relative reduction in gametocyte prevalence by microscopy at day 7 compared to baseline

Diagonal cells indicate percentage relative reduction at day 7 compared to baseline (95% CI) per treatment category, with corresponding p-value compared to baseline. Other cells in the table contain p-values of between-arm comparisons of the relative reductions.

| reference | DHA-PPQ | SP-AQ | PY-AS | AS-AQ | AL | Non-ACT-PQ | ACT-PQ | ACT-TQ |
| --- | --- | --- | --- | --- | --- | --- | --- | --- |
| DHA-PPQ | 19.86% (-14.66% -43.98%, p=0.2258) | p=0.7789 | p=0.9959 | p=0.7557 | p=0.0913 | p=0.0636 | p<0.0001 | p=0.0379 |
| SP-AQ | p=0.7789 | 26.47% (-48.06% -63.48%, p=0.3892) | p=0.8280 | p=0.6201 | p=0.2246 | p=0.1101 | p<0.0001 | p=0.0969 |
| PY-AS | p=0.9959 | p=0.8280 | 20.00% (-73.84% -63.18%, p=0.5731) | p=0.7900 | p=0.1916 | p=0.0945 | p<0.0001 | p=0.0858 |
| AS-AQ | p=0.7557 | p=0.6201 | p=0.7900 | 10.00% (-103.06% -60.11%, p=0.7997) | p=0.1245 | p=0.0660 | p<0.0001 | p=0.0556 |
| AL | p=0.0913 | p=0.2246 | p=0.1916 | p=0.1245 | 51.12% (3.85% -75.15%, p=0.0381) | p=0.4036 | p<0.0001 | p=0.5279 |
| Non-ACT-PQ | p=0.0636 | p=0.1101 | p=0.0945 | p=0.0660 | p=0.4036 | 68.31% (10.02% -88.84%, p=0.0309) | p=0.0340 | p=0.7273 |
| ACT-PQ | p<0.0001 | p<0.0001 | p<0.0001 | p<0.0001 | p<0.0001 | p=0.0340 | 90.52% (78.52% -95.81%, p<0.0001) | p=0.0017 |
| ACT-TQ | p=0.0379 | p=0.0969 | p=0.0858 | p=0.0556 | p=0.5279 | p=0.7273 | p=0.0017 | 61.53% (16.07% -82.37%, p=0.0164) |

### Supplementary Table 5. Relative reduction in gametocyte prevalence by microscopy at day 14 compared to baseline

| reference | DHA-PPQ | SP-AQ | PY-AS | AS-AQ | AL | Non-ACT-PQ | ACT-PQ | ACT-TQ |
| --- | --- | --- | --- | --- | --- | --- | --- | --- |
| DHA-PPQ | 44.37% (16.36% -63.00%, p=0.0048) | p=0.7602 | p=0.9864 | p=0.6191 | p=0.0233 | p=0.0281 | p<0.0001 | p=0.0037 |
| SP-AQ | p=0.7602 | 38.45% (-32.58% -71.42%, p=0.2151) | p=0.8227 | p=0.8134 | p=0.0189 | p=0.0233 | p<0.0001 | p=0.0029 |
| PY-AS | p=0.9864 | p=0.8227 | 44.00% (-33.96% -76.59%, p=0.1926) | p=0.6845 | p=0.0634 | p=0.0326 | p<0.0001 | p=0.0081 |
| AS-AQ | p=0.6191 | p=0.8134 | p=0.6845 | 31.71% (-68.92% -72.39%, p=0.4091) | p=0.0267 | p=0.0211 | p<0.0001 | p=0.0036 |
| AL | p=0.0233 | p=0.0189 | p=0.0634 | p=0.0267 | 75.56% (44.56% -89.23%, p=0.0007) | p=0.1672 | p=0.0009 | p=0.1754 |
| Non-ACT-PQ | p=0.0281 | p=0.0233 | p=0.0326 | p=0.0211 | p=0.1672 | 94.40% (54.76% -99.31%, p=0.0069) | p=0.3857 | p=0.5681 |
| ACT-PQ | p<0.0001 | p<0.0001 | p<0.0001 | p<0.0001 | p=0.0009 | p=0.3857 | 98.10% (91.41% -99.58%, p<0.0001) | p=0.0492 |
| ACT-TQ | p=0.0037 | p=0.0029 | p=0.0081 | p=0.0036 | p=0.1754 | p=0.5681 | p=0.0492 | 89.20% (64.90% -96.68%, p=0.0002) |

Diagonal cells indicate percentage relative reduction at day 14 compared to baseline (95% CI) per treatment category, with corresponding p-value compared to baseline. Other cells in the table contain p-values of between-arm comparisons of the relative reductions.

### Supplementary Table 6. Relative reduction in gametocyte prevalence by RT-qPCR at day 2 compared to baseline

| reference | DHA-PPQ | SP-AQ | PY-AS | AS-AQ | AL | Non-ACT-PQ | ACT-PQ | ACT-TQ |
| --- | --- | --- | --- | --- | --- | --- | --- | --- |
| DHA-PPQ | -0.02% (-37.32% -27.14%, p=0.9989) | p=0.9998 | p=0.9987 | p=0.9995 | p=0.9971 | p=0.9983 | p=0.9995 | p=0.9993 |
| SP-AQ | p=0.9998 | -0.02% (-93.77% -48.37%, p=0.9996) | p=0.9987 | p=0.9997 | p=0.9979 | p=0.9983 | p=0.9998 | p=0.9996 |
| PY-AS | p=0.9987 | p=0.9987 | -0.08% (-110.22% -52.35%, p=0.9983) | p=0.9985 | p=0.9967 | p=0.9996 | p=0.9983 | p=0.9983 |
| AS-AQ | p=0.9995 | p=0.9997 | p=0.9985 | 0.00% (-114.87% -53.46%, p>0.9999) | p=0.9986 | p=0.9982 | p=0.9998 | p>0.9999 |
| AL | p=0.9971 | p=0.9979 | p=0.9967 | p=0.9986 | 0.06% (-77.68% -43.79%, p=0.9982) | p=0.9965 | p=0.9973 | p=0.9982 |
| Non-ACT-PQ | p=0.9983 | p=0.9983 | p=0.9996 | p=0.9982 | p=0.9965 | -0.11% (-126.54% -55.76%, p=0.9980) | p=0.9979 | p=0.9980 |
| ACT-PQ | p=0.9995 | p=0.9998 | p=0.9983 | p=0.9998 | p=0.9973 | p=0.9979 | -0.01% (-66.31% -39.86%, p=0.9997) | p=0.9997 |
| ACT-TQ | p=0.9993 | p=0.9996 | p=0.9983 | p>0.9999 | p=0.9982 | p=0.9980 | p=0.9997 | 0.00% ( -87.91% -46.78%, p>0.9999) |

Diagonal cells indicate percentage relative reduction at day 2 compared to baseline (95% CI) per treatment category, with corresponding p-value compared to baseline. Other cells in the table contain p-values of between-arm comparisons of the relative reductions.

### Supplementary Table 7. Relative reduction in gametocyte prevalence by RT-qPCR at day 7 compared to baseline

Diagonal cells indicate percentage relative reduction at day 7 compared to baseline (95% CI) per treatment category, with corresponding p-value compared to baseline. Other cells in the table contain p-values of between-arm comparisons of the relative reductions.

| reference | DHA-PPQ | SP-AQ | PY-AS | AS-AQ | AL | Non-ACT-PQ | ACT-PQ | ACT-TQ |
| --- | --- | --- | --- | --- | --- | --- | --- | --- |
| DHA-PPQ | 0.08% ( -38.11% -27.71%, p=0.9960) | p=0.9957 | p=0.9972 | p=0.9981 | p=0.9991 | p=0.9970 | p=0.0123 | p=0.9987 |
| SP-AQ | p=0.9957 | -0.08% ( -95.15% -48.68%, p=0.9982) | p=0.9942 | p=0.9985 | p=0.9952 | p=0.9998 | p=0.0532 | p=0.9973 |
| PY-AS | p=0.9972 | p=0.9942 | 0.20% (-109.67% -52.50%, p=0.9957) | p=0.9962 | p=0.9979 | p=0.9953 | p=0.0947 | p=0.9965 |
| AS-AQ | p=0.9981 | p=0.9985 | p=0.9962 | 0.00% (-116.09% -53.72%, p>0.9999) | p=0.9976 | p=0.9989 | p=0.1096 | p=0.9992 |
| AL | p=0.9991 | p=0.9952 | p=0.9979 | p=0.9976 | 0.11% ( -79.11% -44.29%, p=0.9970) | p=0.9965 | p=0.0191 | p=0.9980 |
| Non-ACT-PQ | p=0.9970 | p=0.9998 | p=0.9953 | p=0.9989 | p=0.9965 | -0.06% (-127.64% -56.02%, p=0.9988) | p=0.1393 | p=0.9980 |
| ACT-PQ | p=0.0123 | p=0.0532 | p=0.0947 | p=0.1096 | p=0.0191 | p=0.1393 | 42.91% (1.55% -66.89%, p=0.0438) | p=0.0414 |
| ACT-TQ | p=0.9987 | p=0.9973 | p=0.9965 | p=0.9992 | p=0.9980 | p=0.9980 | p=0.0414 | 0.04% ( -89.91% -47.38%, p=0.9991) |

### Supplementary Table 8. Relative reduction in gametocyte prevalence by RT-qPCR at day 14 compared to baseline

Diagonal cells indicate percentage relative reduction at day 14 compared to baseline (95% CI) per treatment category, with corresponding p-value compared to baseline. Other cells in the table contain p-values of between-arm comparisons of the relative reductions.

| reference | DHA-PPQ | SP-AQ | PY-AS | AS-AQ | AL | Non-ACT-PQ | ACT-PQ | ACT-TQ |
| --- | --- | --- | --- | --- | --- | --- | --- | --- |
| DHA-PPQ | 0.15% (-37.69% -27.59%, p=0.9926) | p=0.9939 | p=0.8986 | p=0.8834 | p=0.8295 | p=0.1125 | p<0.0001 | p=0.3296 |
| SP-AQ | p=0.9939 | -0.08% (-94.71% -48.56%, p=0.9982) | p=0.9062 | p=0.8917 | p=0.8573 | p=0.1380 | p<0.0001 | p=0.4032 |
| PY-AS | p=0.8986 | p=0.9062 | 4.43% (-101.51% -54.67%, p=0.9053) | p=0.9827 | p=0.9778 | p=0.1863 | p=0.0007 | p=0.5252 |
| AS-AQ | p=0.8834 | p=0.8917 | p=0.9827 | 5.34% (-106.97% -56.71%, p=0.8906) | p=0.9994 | p=0.2031 | p=0.0015 | p=0.5590 |
| AL | p=0.8295 | p=0.8573 | p=0.9778 | p=0.9994 | 5.37% (-69.78% -47.25%, p=0.8532) | p=0.1474 | p<0.0001 | p=0.4455 |
| Non-ACT-PQ | p=0.1125 | p=0.1380 | p=0.1863 | p=0.2031 | p=0.1474 | 53.25% (-25.90% -82.64%, p=0.1325) | p=0.3048 | p=0.3673 |
| ACT-PQ | p<0.0001 | p<0.0001 | p=0.0007 | p=0.0015 | p<0.0001 | p=0.3048 | 71.76% (48.60% -84.48%, p<0.0001) | p=0.0026 |
| ACT-TQ | p=0.3296 | p=0.4032 | p=0.5252 | p=0.5590 | p=0.4455 | p=0.3673 | p=0.0026 | 25.59% (-45.84% -62.03%, p=0.3893) |

### Supplementary Figure 4. Relative reduction in gametocyte prevalence per study arm (ungrouped)

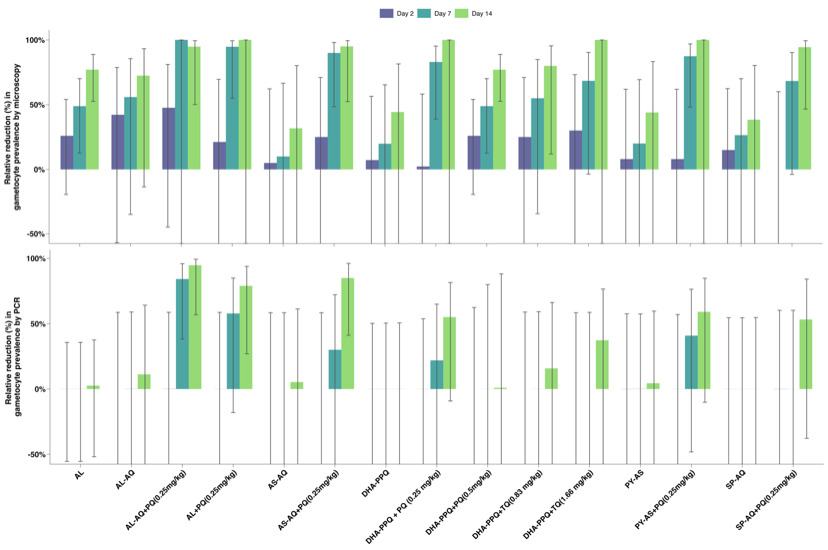

### Supplementary Table 9. Relative reduction in gametocyte density by microscopy at day 2 compared to baseline

Diagonal cells indicate percentage relative reduction at day 2 compared to baseline (95% CI) per treatment category, with corresponding p-value compared to baseline. Other cells in the table contain p-values of between-arm comparisons of the relative reductions.

| reference | | DHA-PPQ | SP-AQ | PY-AS | AS-AQ | AL | Non-ACT-PQ | ACT-PQ | ACT-TQ |
| --- | --- | --- | --- | --- | --- | --- | --- | --- | --- |
| DHA-PPQ | 18.59% (-49.99% -55.81%, p=0.5094) | | p=0.2270 | p=0.3077 | p=0.9348 | p=0.0489 | p=0.3718 | p=0.1502 | p=0.2895 |
| SP-AQ | p=0.2270 | | -23.83% (-150.43% -38.77%, p=0.5519) | p=0.0607 | p=0.3406 | p=0.0040 | p=0.9612 | p=0.0124 | p=0.0439 |
| PY-AS | p=0.3077 | | p=0.0607 | 46.29% (-21.88% -76.33%, p=0.1371) | p=0.4686 | p=0.6389 | p=0.1212 | p=0.9317 | p=0.9138 |
| AS-AQ | p=0.9348 | | p=0.3406 | p=0.4686 | 21.49% (-90.80% -67.70%, p=0.5933) | p=0.2033 | p=0.4335 | p=0.4125 | p=0.4882 |
| AL | p=0.0489 | | p=0.0040 | p=0.6389 | p=0.2033 | 55.84% (16.68% -76.59%, p=0.0116) | p=0.0260 | p=0.4143 | p=0.4958 |
| Non-ACT-PQ | p=0.3718 | | p=0.9612 | p=0.1212 | p=0.4335 | p=0.0260 | -20.98% (-194.03% -50.22%, p=0.6742) | p=0.0657 | p=0.1103 |
| ACT-PQ | p=0.1502 | | p=0.0124 | p=0.9317 | p=0.4125 | p=0.4143 | p=0.0657 | 44.49% (23.64% -59.64%, p=0.0003) | p=0.9622 |
| ACT-TQ | p=0.2895 | | p=0.0439 | p=0.9138 | p=0.4882 | p=0.4958 | p=0.1103 | p=0.9622 | 43.64% (-13.98% -72.13%, p=0.1105) |

### Supplementary Table 10. Relative reduction in gametocyte density by microscopy at day 7 compared to baseline

Diagonal cells indicate percentage relative reduction at day 7 compared to baseline (95% CI) per treatment category, with corresponding p-value compared to baseline. Other cells in the table contain p-values of between-arm comparisons of the relative reductions.

| reference | | DHA-PPQ | SP-AQ | PY-AS | AS-AQ | AL | Non-ACT-PQ | ACT-PQ | ACT-TQ |
| --- | --- | --- | --- | --- | --- | --- | --- | --- | --- |
| DHA-PPQ | 52.44% (12.02% -74.29%, p=0.0179) | | p=0.1550 | p=0.4917 | p=0.7638 | p=0.0015 | p<0.0001 | p<0.0001 | p<0.0001 |
| SP-AQ | p=0.1550 | | 21.53% (-59.92% -61.49%, p=0.5045) | p=0.0810 | p=0.4450 | p<0.0001 | p<0.0001 | p<0.0001 | p<0.0001 |
| PY-AS | p=0.4917 | | p=0.0810 | 64.10% (18.46% -84.19%, p=0.0144) | p=0.4285 | p=0.0872 | p=0.0002 | p<0.0001 | p=0.0018 |
| AS-AQ | p=0.7638 | | p=0.4450 | p=0.4285 | 45.64% (-32.20% -77.65%, p=0.1788) | p=0.0127 | p<0.0001 | p<0.0001 | p=0.0002 |
| AL | p=0.0015 | | p<0.0001 | p=0.0872 | p=0.0127 | 82.43% (66.76% -90.72%, p<0.0001) | p=0.0056 | p<0.0001 | p=0.0591 |
| Non-ACT-PQ | p<0.0001 | | p<0.0001 | p=0.0002 | p<0.0001 | p=0.0056 | 95.08% (87.89% -98.00%, p<0.0001) | p<0.0001 | p=0.2228 |
| ACT-PQ | p<0.0001 | | p<0.0001 | p<0.0001 | p<0.0001 | p<0.0001 | p<0.0001 | 99.64% (99.50% -99.74%, p<0.0001) | p<0.0001 |
| ACT-TQ | p<0.0001 | | p<0.0001 | p=0.0018 | p=0.0002 | p=0.0591 | p=0.2228 | p<0.0001 | 91.11% (81.94% -95.62%, p<0.0001) |

### Supplementary Table 11. Relative reduction in gametocyte density by microscopy at day 14 compared to baseline

| reference | DHA-PPQ | SP-AQ | PY-AS | AS-AQ | AL | Non-ACT-PQ | ACT-PQ | ACT-TQ |
| --- | --- | --- | --- | --- | --- | --- | --- | --- |
| DHA-PPQ | 79.32% (61.63% -88.85%, p<0.0001) | p=0.1648 | p=0.6312 | p=0.5357 | p<0.0001 | p<0.0001 | p<0.0001 | p<0.0001 |
| SP-AQ | p=0.1648 | 66.29% (31.53% -83.40%, p=0.0026) | p=0.1249 | p=0.6658 | p<0.0001 | p<0.0001 | p<0.0001 | p<0.0001 |
| PY-AS | p=0.6312 | p=0.1249 | 83.01% (61.42% -92.52%, p<0.0001) | p=0.3675 | p=0.0007 | p<0.0001 | p<0.0001 | p<0.0001 |
| AS-AQ | p=0.5357 | p=0.6658 | p=0.3675 | 72.65% (32.79% -88.87%, p=0.0047) | p<0.0001 | p<0.0001 | p<0.0001 | p<0.0001 |
| AL | p<0.0001 | p<0.0001 | p=0.0007 | p<0.0001 | 95.88% (92.20% -97.82%, p<0.0001) | p<0.0001 | p<0.0001 | p<0.0001 |
| Non-ACT-PQ | p<0.0001 | p<0.0001 | p<0.0001 | p<0.0001 | p<0.0001 | 99.70% (99.25% -99.88%, p<0.0001) | p=0.1110 | p=0.0778 |
| ACT-PQ | p<0.0001 | p<0.0001 | p<0.0001 | p<0.0001 | p<0.0001 | p=0.1110 | 99.85% (99.79% -99.89%, p<0.0001) | p<0.0001 |
| ACT-TQ | p<0.0001 | p<0.0001 | p<0.0001 | p<0.0001 | p<0.0001 | p=0.0778 | p<0.0001 | 99.28% (98.53% -99.65%, p<0.0001) |

Diagonal cells indicate percentage relative reduction at day 14 compared to baseline (95% CI) per treatment category, with corresponding p-value compared to baseline. Other cells in the table contain p-values of between-arm comparisons of the relative reductions.

### Supplementary Table 12. Relative reduction in gametocyte density by RT-qPCR at day 2 compared to baseline

| reference | DHA-PPQ | SP-AQ | PY-AS | AS-AQ | AL | Non-ACT-PQ | ACT-PQ | ACT-TQ |
| --- | --- | --- | --- | --- | --- | --- | --- | --- |
| DHA-PPQ | 21.48% (-14.69% -46.25%, p=0.2109) | p=0.8363 | p=0.8914 | p=0.7188 | p=0.0036 | p=0.7777 | p<0.0001 | p=0.6758 |
| SP-AQ | p=0.8363 | 25.22% (-20.10% -53.44%, p=0.2292) | p=0.7818 | p=0.8642 | p=0.0308 | p=0.9113 | p=0.0035 | p=0.8649 |
| PY-AS | p=0.8914 | p=0.7818 | 18.46% (-41.54% -53.03%, p=0.4682) | p=0.6888 | p=0.0302 | p=0.7359 | p=0.0056 | p=0.6644 |
| AS-AQ | p=0.7188 | p=0.8642 | p=0.6888 | 29.25% (-25.84% -60.22%, p=0.2390) | p=0.1085 | p=0.9630 | p=0.0335 | p=0.9756 |
| AL | p=0.0036 | p=0.0308 | p=0.0302 | p=0.1085 | 56.18% (34.26% -70.79%, p<0.0001) | p=0.1154 | p=0.5477 | p=0.0398 |
| Non-ACT-PQ | p=0.7777 | p=0.9113 | p=0.7359 | p=0.9630 | p=0.1154 | 27.99% (-32.47% -60.86%, p=0.2910) | p=0.0400 | p=0.9811 |
| ACT-PQ | p<0.0001 | p=0.0035 | p=0.0056 | p=0.0335 | p=0.5477 | p=0.0400 | 60.70% (52.17% -67.71%, p<0.0001) | p=0.0043 |
| ACT-TQ | p=0.6758 | p=0.8649 | p=0.6644 | p=0.9756 | p=0.0398 | p=0.9811 | p=0.0043 | 28.56% (-12.53% -54.65%, p=0.1469) |

### Supplementary Table 13. Relative reduction in gametocyte density by RT-qPCR at day 7 compared to baseline

Diagonal cells indicate percentage relative reduction at day 2 compared to baseline (95% CI) per treatment category, with corresponding p-value compared to baseline. Other cells in the table contain p-values of between-arm comparisons of the relative reductions.

| reference | DHA-PPQ | SP-AQ | PY-AS | AS-AQ | AL | Non-ACT-PQ | ACT-PQ | ACT-TQ |
| --- | --- | --- | --- | --- | --- | --- | --- | --- |
| DHA-PPQ | 55.14% (33.47% -69.75%, p<0.0001) | p=0.5973 | p=0.9593 | p=0.8502 | p=0.0026 | p<0.0001 | p<0.0001 | p=0.0001 |
| SP-AQ | p=0.5973 | 49.12% (17.61% -68.58%, p=0.0060) | p=0.7179 | p=0.5761 | p=0.0029 | p<0.0001 | p<0.0001 | p=0.0002 |
| PY-AS | p=0.9593 | p=0.7179 | 54.51% (20.93% -73.83%, p=0.0052) | p=0.8445 | p=0.0271 | p<0.0001 | p<0.0001 | p=0.0029 |
| AS-AQ | p=0.8502 | p=0.5761 | p=0.8445 | 57.54% (23.99% -76.28%, p=0.0039) | p=0.0617 | p<0.0001 | p<0.0001 | p=0.0085 |
| AL | p=0.0026 | p=0.0029 | p=0.0271 | p=0.0617 | 75.71% (63.18% -83.98%, p<0.0001) | p<0.0001 | p<0.0001 | p=0.2454 |
| Non-ACT-PQ | p<0.0001 | p<0.0001 | p<0.0001 | p<0.0001 | p<0.0001 | 98.87% (97.91% -99.39%, p<0.0001) | p<0.0001 | p<0.0001 |
| ACT-PQ | p<0.0001 | p<0.0001 | p<0.0001 | p<0.0001 | p<0.0001 | p<0.0001 | 99.67% (99.60% -99.73%, p<0.0001) | p<0.0001 |
| ACT-TQ | p=0.0001 | p=0.0002 | p=0.0029 | p=0.0085 | p=0.2454 | p<0.0001 | p<0.0001 | 81.64% (70.69% -88.50%, p<0.0001) |

Diagonal cells indicate percentage relative reduction at day 7 compared to baseline (95% CI) per treatment category, with corresponding p-value compared to baseline. Other cells in the table contain p-values of between-arm comparisons of the relative reductions.

### Supplementary Table 14. Relative reduction in gametocyte density by RT-qPCR at day 14 compared to baseline

| reference | DHA-PPQ | SP-AQ | PY-AS | AS-AQ | AL | Non-ACT-PQ | ACT-PQ | ACT-TQ |
| --- | --- | --- | --- | --- | --- | --- | --- | --- |
| DHA-PPQ | 71.75% (58.07% -80.96%, p<0.0001) | p=0.8226 | p=0.9919 | p=0.4020 | p<0.0001 | p<0.0001 | p<0.0001 | p<0.0001 |
| SP-AQ | p=0.8226 | 70.20% (51.58% -81.65%, p<0.0001) | p=0.8705 | p=0.3599 | p<0.0001 | p<0.0001 | p<0.0001 | p<0.0001 |
| PY-AS | p=0.9919 | p=0.8705 | 71.67% (50.66% -83.73%, p<0.0001) | p=0.4825 | p<0.0001 | p<0.0001 | p<0.0001 | p<0.0001 |
| AS-AQ | p=0.4020 | p=0.3599 | p=0.4825 | 77.92% (60.11% -87.78%, p<0.0001) | p=0.0013 | p<0.0001 | p<0.0001 | p<0.0001 |
| AL | p<0.0001 | p<0.0001 | p<0.0001 | p=0.0013 | 91.67% (87.34% -94.52%, p<0.0001) | p<0.0001 | p<0.0001 | p<0.0001 |
| Non-ACT-PQ | p<0.0001 | p<0.0001 | p<0.0001 | p<0.0001 | p<0.0001 | 99.97% (99.95% -99.99%, p<0.0001) | p=0.0020 | p<0.0001 |
| ACT-PQ | p<0.0001 | p<0.0001 | p<0.0001 | p<0.0001 | p<0.0001 | p=0.0020 | 99.94% (99.92% -99.95%, p<0.0001) | p<0.0001 |
| ACT-TQ | p<0.0001 | p<0.0001 | p<0.0001 | p<0.0001 | p<0.0001 | p<0.0001 | p<0.0001 | 99.10% (98.56% -99.44%, p<0.0001) |

Diagonal cells indicate percentage relative reduction at day 14 compared to baseline (95% CI) per treatment category, with corresponding p-value compared to baseline. Other cells in the table contain p-values of between-arm comparisons of the relative reductions.

### Supplementary Figure 5. Relative reduction in gametocyte densities per study arm (ungrouped)

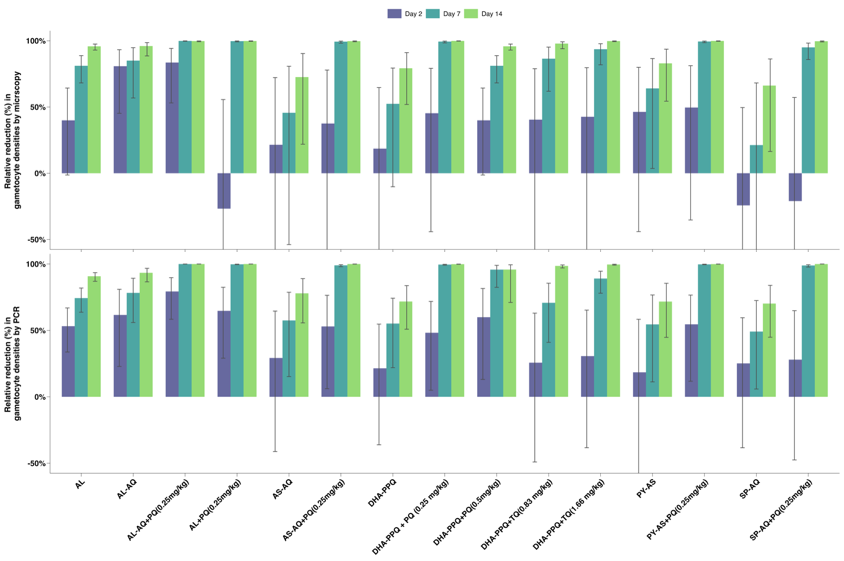

### Supplementary Table 15. Relative reduction in mosquito infection rate at day 2 compared to baseline

| reference | DHA-PPQ | SP-AQ | PY-AS | AS-AQ | AL | Non-ACT-PQ | ACT-PQ | ACT-TQ |
| --- | --- | --- | --- | --- | --- | --- | --- | --- |
| DHA-PPQ | 20.35% (12.70% -27.33%, p<0.0001) | p=0.0002 | p=0.0009 | p<0.0001 | p<0.0001 | p<0.0001 | p<0.0001 | p<0.0001 |
| SP-AQ | p=0.0002 | 2.17% (-12.80% -15.14%, p=0.7629) | p=0.3360 | p=0.0259 | p<0.0001 | p<0.0001 | p<0.0001 | p=0.2729 |
| PY-AS | p=0.0009 | p=0.3360 | -5.18% (-26.88% -12.81%, p=0.5974) | p=0.2873 | p<0.0001 | p<0.0001 | p<0.0001 | p=0.8795 |
| AS-AQ | p<0.0001 | p=0.0259 | p=0.2873 | -17.32% (-42.92% - 3.69%, p=0.1126) | p<0.0001 | p<0.0001 | p<0.0001 | p=0.1693 |
| AL | p<0.0001 | p<0.0001 | p<0.0001 | p<0.0001 | 97.10% (94.84% -98.37%, p<0.0001) | p<0.0001 | p=0.0097 | p<0.0001 |
| Non-ACT-PQ | p<0.0001 | p<0.0001 | p<0.0001 | p<0.0001 | p<0.0001 | 83.26% (78.11% -87.21%, p<0.0001) | p<0.0001 | p<0.0001 |
| ACT-PQ | p<0.0001 | p<0.0001 | p<0.0001 | p<0.0001 | p=0.0097 | p<0.0001 | 98.95% (98.19% -99.39%, p<0.0001) | p<0.0001 |
| ACT-TQ | p<0.0001 | p=0.2729 | p=0.8795 | p=0.1693 | p<0.0001 | p<0.0001 | p<0.0001 | -3.87% (-21.61% -11.28%, p=0.6369) |

Diagonal cells indicate percentage relative reduction at day 2 compared to baseline (95% CI) per treatment category, with corresponding p-value compared to baseline. Other cells in the table contain p-values of between-arm comparisons of the relative reductions.

### Supplementary Table 16. Relative reduction in mosquito infection rate at day 7 compared to baseline

| reference | DHA-PPQ | SP-AQ | PY-AS | AS-AQ | AL | Non-ACT-PQ | ACT-PQ | ACT-TQ |
| --- | --- | --- | --- | --- | --- | --- | --- | --- |
| DHA-PPQ | 41.51% (34.98% - 47.37%, p<0.0001) | p<0.0001 | p<0.0001 | p=0.0678 | p=0.0001 | p=0.0045 | p<0.0001 | p<0.0001 |
| SP-AQ | p<0.0001 | 9.79% (-6.54% - 23.61%, p=0.2249) | p=0.7647 | p<0.0001 | p<0.0001 | p=0.0026 | p<0.0001 | p<0.0001 |
| PY-AS | p<0.0001 | p=0.7647 | 7.58% (-13.93% - 24.73%, p=0.4517) | p<0.0001 | p<0.0001 | p=0.0025 | p<0.0001 | p<0.0001 |
| AS-AQ | p=0.0536 | p<0.0001 | p<0.0001 | 55.32% (39.21% - 67.16%, p<0.0001) | p=0.0002 | p=0.0062 | p<0.0001 | p<0.0001 |
| AL | p=1e-04 | p<0.0001 | p<0.0001 | p=2e-04 | 99.92% (97.63% -100.00%, p<0.0001) | p=0.8048 | p=0.4754 | p=0.2680 |
| Non-ACT-PQ | p=0.0044 | p=0.0026 | p=0.0025 | p=0.0062 | p=0.8048 | 99.96% (93.98% -100.00%, p=0.0023) | p=0.4379 | p=0.2932 |
| ACT-PQ | p<0.0001 | p<0.0001 | p<0.0001 | p<0.0001 | p=0.4754 | p=0.4379 | 99.70% (99.18% - 99.89%, p<0.0001) | p=0.3685 |
| ACT-TQ | p<0.0001 | p<0.0001 | p<0.0001 | p<0.0001 | p=0.2680 | p=0.2932 | p=0.3685 | 99.37% (97.60% - 99.83%, p<0.0001) |

Diagonal cells indicate percentage relative reduction at day 7 compared to baseline (95% CI) per treatment category, with corresponding p-value compared to baseline. Other cells in the table contain p-values of between-arm comparisons of the relative reductions.

### Supplementary Table 17. Relative reduction in mosquito infection rate at day 14 compared to baseline

| reference | DHA-PPQ | SP-AQ | PY-AS | AS-AQ | AL | Non-ACT-PQ | ACT-PQ | ACT-TQ |
| --- | --- | --- | --- | --- | --- | --- | --- | --- |
| DHA-PPQ | 64.05% (57.68% - 69.46%, p<0.0001) | p=1e-04 | p=0.1469 | p=0.0194 | p=0.0058 | p>0.9999 | p=0.0078 | p=0.0098 |
| SP-AQ | p=1e-04 | 81.65% (73.10% - 87.48%, p<0.0001) | p=0.0320 | p=0.6313 | p=0.0144 | p>0.9999 | p=0.0197 | p=0.0213 |
| PY-AS | p=0.1469 | p=0.0320 | 71.44% (59.42% - 79.91%, p<0.0001) | p=0.2203 | p=0.0081 | p>0.9999 | p=0.0109 | p=0.0129 |
| AS-AQ | p=0.0194 | p=0.6313 | p=0.2203 | 79.14% (66.13% - 87.15%, p<0.0001) | p=0.0125 | p>0.9999 | p=0.0170 | p=0.0187 |
| AL | p=0.0058 | p=0.0144 | p=0.0081 | p=0.0125 | 99.91% (93.71% -100.00%, p=0.0013) | p>0.9999 | p=0.8763 | p=0.9430 |
| Non-ACT-PQ | p=0.0098 | p=0.0213 | p=0.0129 | p=0.0187 | p=0.9430 | 64.05% (57.68% - 69.46%, p<0.0001) | p=0.8259 | p>0.9999 |
| ACT-PQ | p=0.0078 | p=0.0197 | p=0.0109 | p=0.0170 | p=0.8763 | p>0.9999 | 99.86% (91.65% -100.00%, p=0.0017) | p=0.8259 |
| ACT-TQ | p=0.0098 | p=0.0213 | p=0.0129 | p=0.0187 | p=0.9430 | p>0.9999 | p=0.8259 | 99.93% (92.06% -100.00%, p=0.0027) |

Diagonal cells indicate percentage relative reduction at day 14 compared to baseline (95% CI) per treatment category, with corresponding p-value compared to baseline. Other cells in the table contain p-values of between-arm comparisons of the relative reductions.

### Supplementary Figure 6. Relative reduction in mosquito infection rate per study arm (ungrouped)

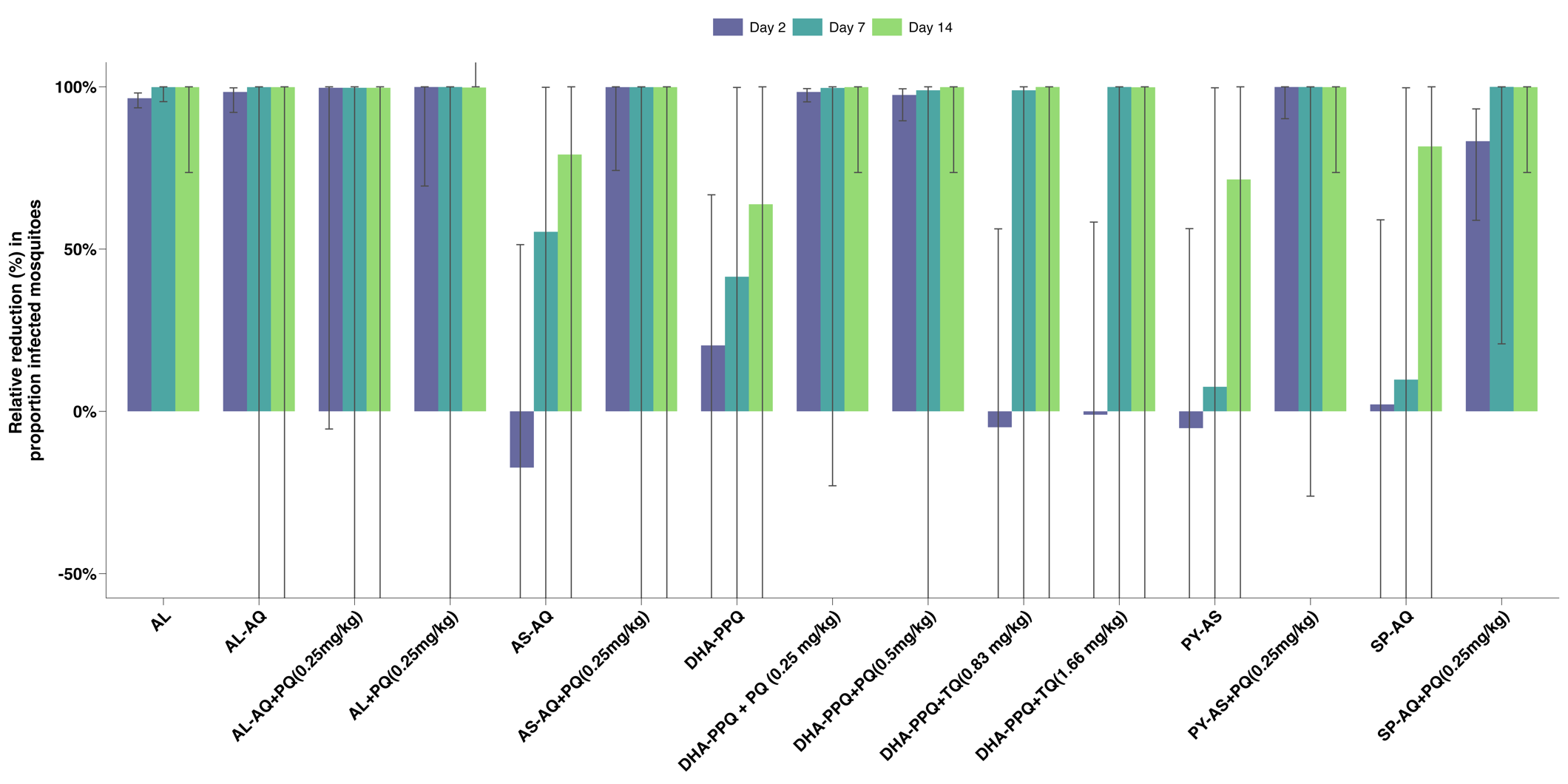

### Supplementary Figure 7. Relative reduction in mosquito infection rate comparing the same study arms across different studies

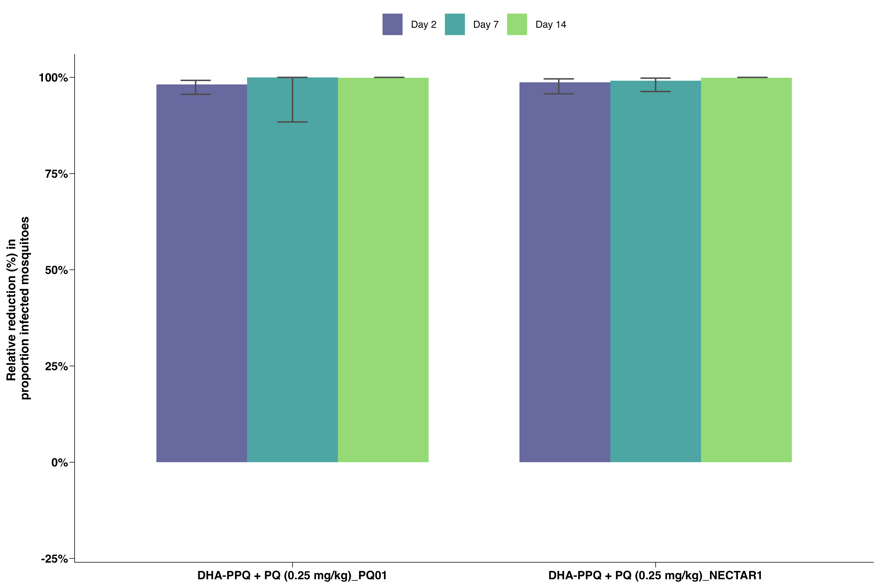

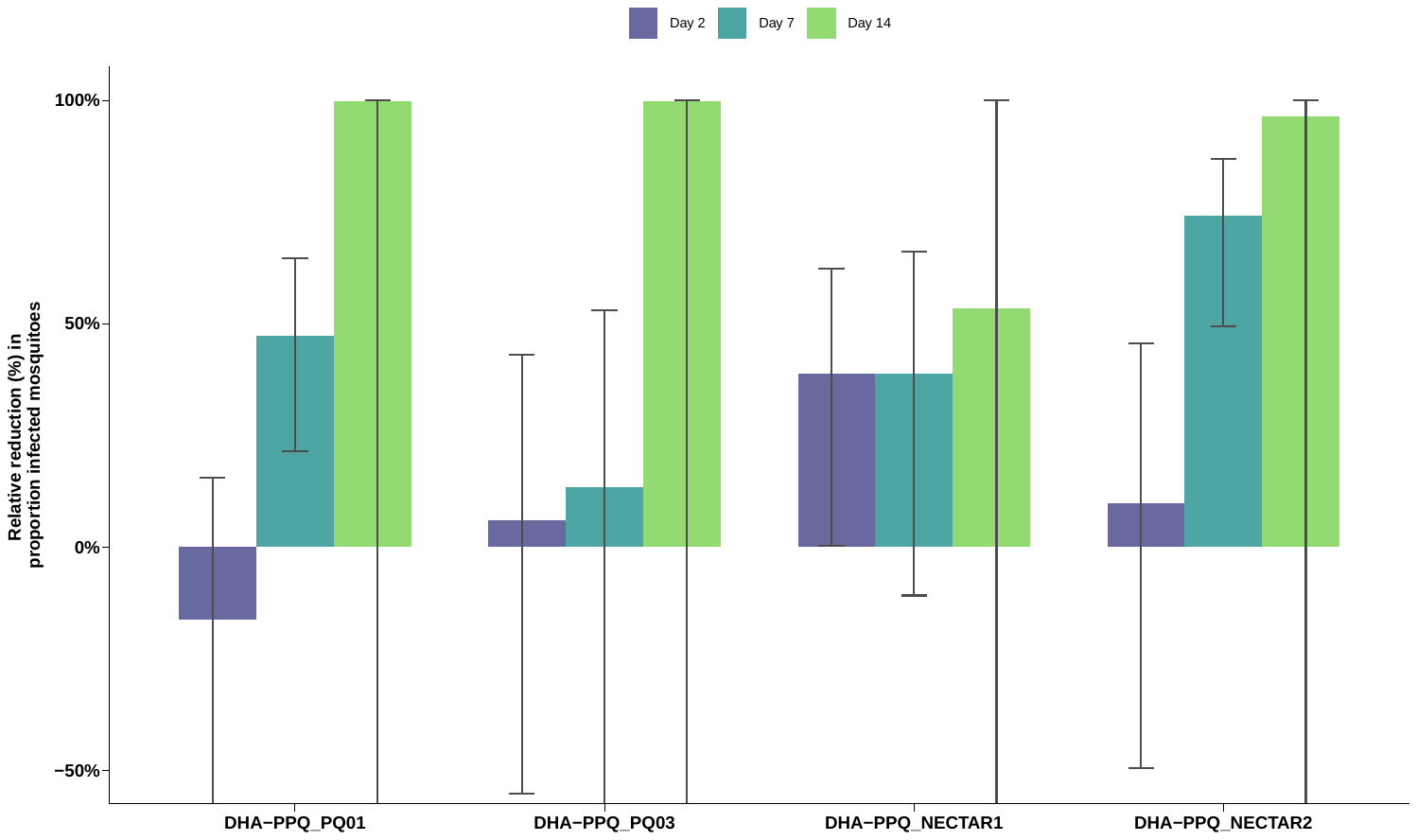

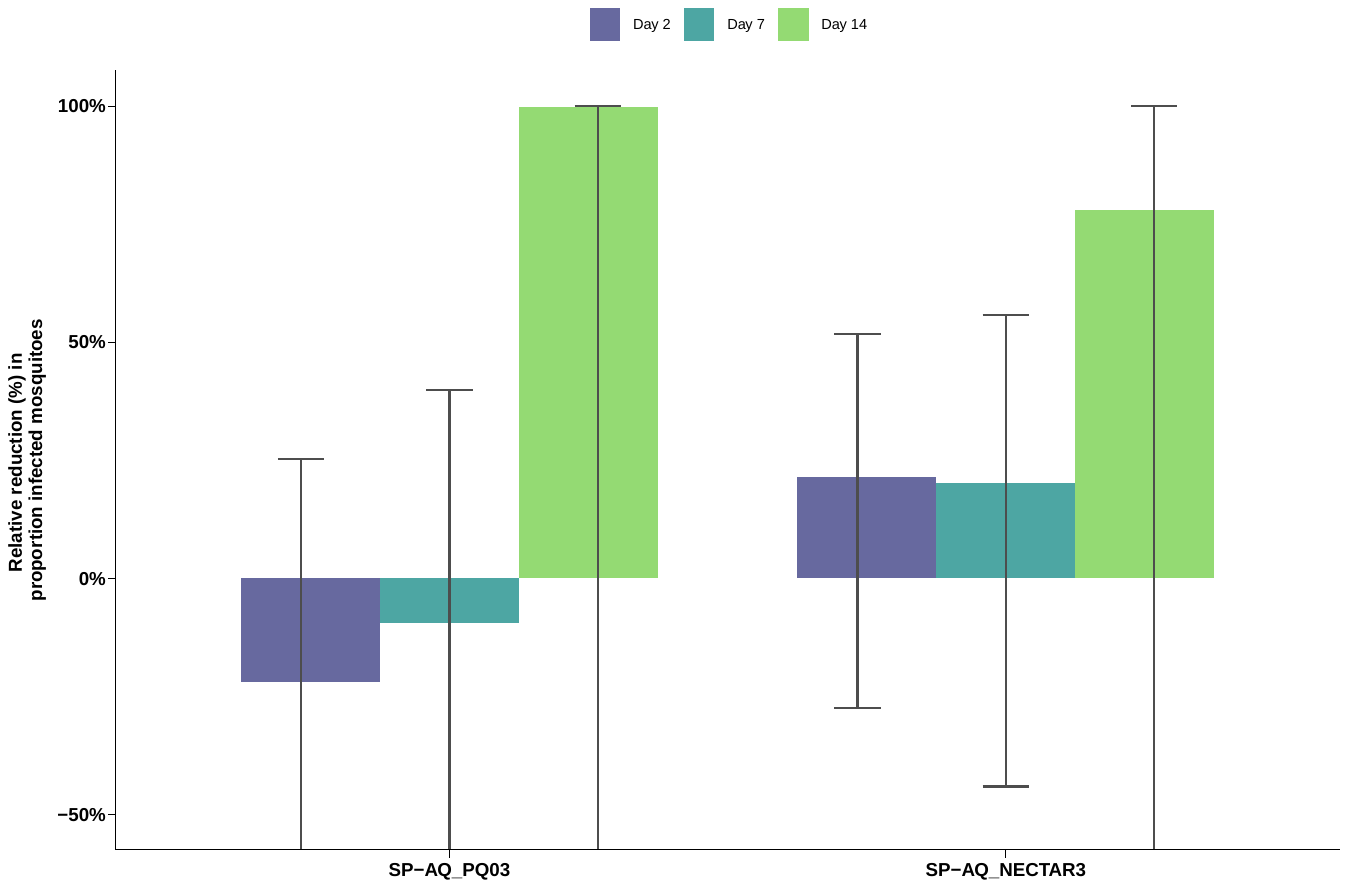

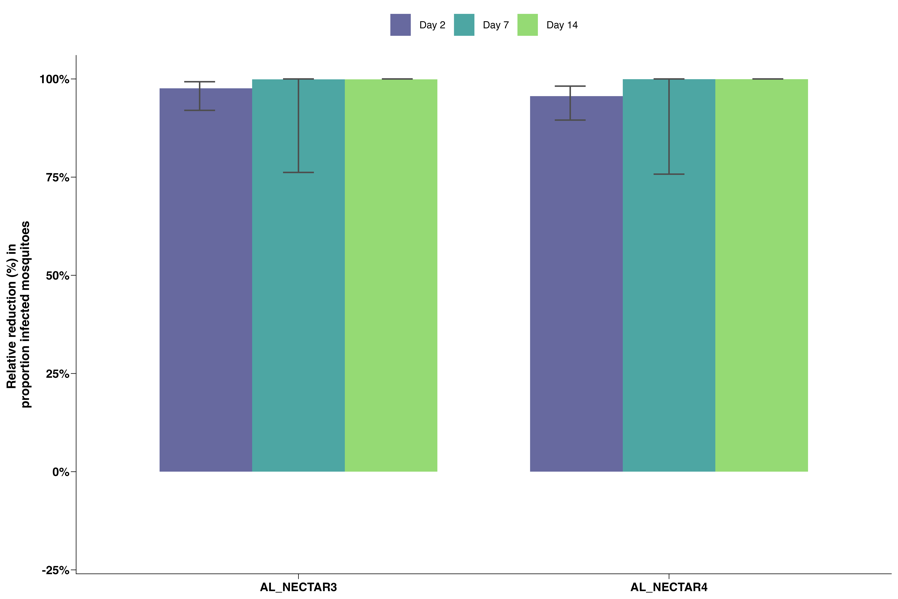

### Supplementary Figure 8. Relative reduction in the probability of infecting at least 1 mosquito

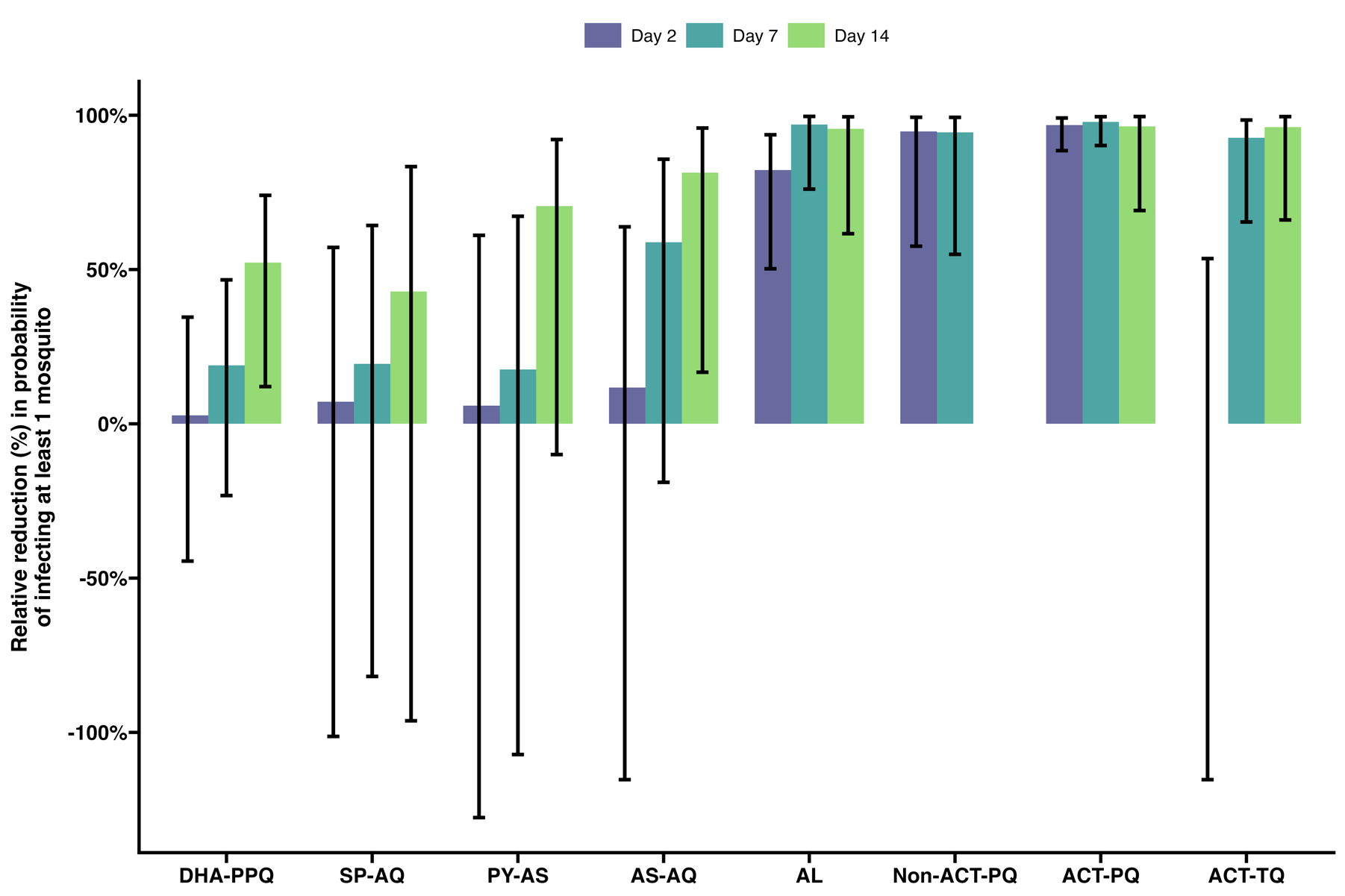

### Supplementary Table 18. Relative reduction in the probability of infecting at least 1 mosquito at day 2 compared to baseline

| reference | DHA-PPQ | SP-AQ | PY-AS | AS-AQ | AL | Non-ACT-PQ | ACT-PQ | ACT-TQ |
| --- | --- | --- | --- | --- | --- | --- | --- | --- |
| DHA-PPQ | 2.77% (-44.48% -34.57%, p=0.8895) | p=0.8920 | p=0.9356 | p=0.8118 | p=0.0005 | p=0.0053 | p<0.0001 | p=0.9332 |
| SP-AQ | p=0.8920 | 7.14% (-101.30% -57.17%, p=0.8511) | p=0.9757 | p=0.9090 | p=0.0014 | p=0.0069 | p<0.0001 | p=0.8460 |
| PY-AS | p=0.9356 | p=0.9757 | 5.88% (-127.61% -61.08%, p=0.8930) | p=0.8966 | p=0.0030 | p=0.0078 | p<0.0001 | p=0.8902 |
| AS-AQ | p=0.8118 | p=0.9090 | p=0.8966 | 11.76% (-115.31% -63.84%, p=0.7833) | p=0.0046 | p=0.0094 | p<0.0001 | p=0.7779 |
| AL | p=0.0005 | p=0.0014 | p=0.0030 | p=0.0046 | 82.27% (50.26% -93.68%, p=0.0010) | p=0.2768 | p=0.0196 | p=0.0008 |
| Non-ACT-PQ | p=0.0053 | p=0.0069 | p=0.0078 | p=0.0094 | p=0.2768 | 94.74% (57.56% -99.35%, p=0.0057) | p=0.6726 | p=0.0055 |
| ACT-PQ | p<0.0001 | p<0.0001 | p<0.0001 | p<0.0001 | p=0.0196 | p=0.6726 | 96.81% (88.53% -99.11%, p<0.0001) | p<0.0001 |
| ACT-TQ | p=0.9332 | p=0.8460 | p=0.8902 | p=0.7779 | p=0.0008 | p=0.0055 | p<0.0001 | 0.00% (-115.31% -53.55%, p>0.9999) |

Diagonal cells indicate percentage relative reduction at day 2 compared to baseline (95% CI) per treatment category, with corresponding p-value compared to baseline. Other cells in the table contain p-values of between-arm comparisons of the relative reductions.

### Supplementary Table 19. Relative reduction in the probability of infecting at least 1 mosquito at day 7 compared to baseline

| reference | DHA-PPQ | SP-AQ | PY-AS | AS-AQ | AL | Non-ACT-PQ | ACT-PQ | ACT-TQ |
| --- | --- | --- | --- | --- | --- | --- | --- | --- |
| DHA-PPQ | 18.92% (-23.24% -46.66%, p=0.3262) | p=0.9863 | p=0.9703 | p=0.1731 | p=0.0015 | p=0.0105 | p<0.0001 | p=0.0016 |
| SP-AQ | p=0.9863 | 19.41% (-81.86% -64.29%, p=0.6032) | p=0.9624 | p=0.2068 | p=0.0018 | p=0.0119 | p<0.0001 | p=0.0023 |
| PY-AS | p=0.9703 | p=0.9624 | 17.65% (-107.17% -67.26%, p=0.6800) | p=0.2289 | p=0.0021 | p=0.0131 | p<0.0001 | p=0.0030 |
| AS-AQ | p=0.1731 | p=0.2068 | p=0.2289 | 58.82% (-18.95% -85.75%, p=0.1011) | p=0.0183 | p=0.0733 | p=0.0005 | p=0.0444 |
| AL | p=0.0015 | p=0.0018 | p=0.0021 | p=0.0183 | 96.99% (76.07% -99.62%, p=0.0009) | p=0.6720 | p=0.7854 | p=0.4766 |
| Non-ACT-PQ | p=0.0105 | p=0.0119 | p=0.0131 | p=0.0733 | p=0.6720 | 94.46% (54.93% -99.32%, p=0.0068) | p=0.4480 | p=0.8245 |
| ACT-PQ | p<0.0001 | p<0.0001 | p<0.0001 | p=0.0005 | p=0.7854 | p=0.4480 | 97.85% (90.19% -99.53%, p<0.0001) | p=0.2299 |
| ACT-TQ | p=0.0016 | p=0.0023 | p=0.0030 | p=0.0444 | p=0.4766 | p=0.8245 | p=0.2299 | 92.67% (65.42% -98.45%, p=0.0010) |

Diagonal cells indicate percentage relative reduction at day 7 compared to baseline (95% CI) per treatment category, with corresponding p-value compared to baseline. Other cells in the table contain p-values of between-arm comparisons of the relative reductions.

### Supplementary Table 20. Relative reduction in the probability of infecting at least 1 mosquito at day 14 compared to baseline

| reference | DHA-PPQ | SP-AQ | PY-AS | AS-AQ | AL | Non-ACT-PQ | ACT-PQ | ACT-TQ |
| --- | --- | --- | --- | --- | --- | --- | --- | --- |
| DHA-PPQ | 52.23% (12.07% -74.05%, p=0.0176) | p=0.7432 | p=0.4162 | p=0.1768 | p=0.0245 | p>0.9999 | p=0.0139 | p=0.0181 |
| SP-AQ | p=0.7432 | 42.86% (-96.23% -83.36%, p=0.3740) | p=0.3281 | p=0.1450 | p=0.0208 | p>0.9999 | p=0.0120 | p=0.0154 |
| PY-AS | p=0.4162 | p=0.3281 | 70.59% (-9.96% -92.13%, p=0.0689) | p=0.5690 | p=0.0939 | p>0.9999 | p=0.0620 | p=0.0743 |
| AS-AQ | p=0.1768 | p=0.1450 | p=0.5690 | 81.42% (16.72% -95.86%, p=0.0279) | p=0.2266 | p>0.9999 | p=0.1654 | p=0.1886 |
| AL | p=0.0245 | p=0.0208 | p=0.0939 | p=0.2266 | 95.60% (61.62% -99.50%, p=0.0047) | p>0.9999 | p=0.8878 | p=0.9281 |
| Non-ACT-PQ | p=0.0181 | p=0.0154 | p=0.0743 | p=0.1886 | p=0.9281 | 52.23% (12.07% -74.05%, p=0.0176) | p=0.9600 | p>0.9999 |
| ACT-PQ | p=0.0139 | p=0.0120 | p=0.0620 | p=0.1654 | p=0.8878 | p>0.9999 | 96.41% (69.12% -99.58%, p=0.0024) | p=0.9600 |
| ACT-TQ | p=0.0181 | p=0.0154 | p=0.0743 | p=0.1886 | p=0.9281 | p>0.9999 | p=0.9600 | 96.14% (66.07% -99.56%, p=0.0033) |

Diagonal cells indicate percentage relative reduction at day 14 compared to baseline (95% CI) per treatment category, with corresponding p-value compared to baseline. Other cells in the table contain p-values of between-arm comparisons of the relative reductions.

### Supplementary Figure 9. Relative reduction in mean oocyst density in dissected mosquitoes

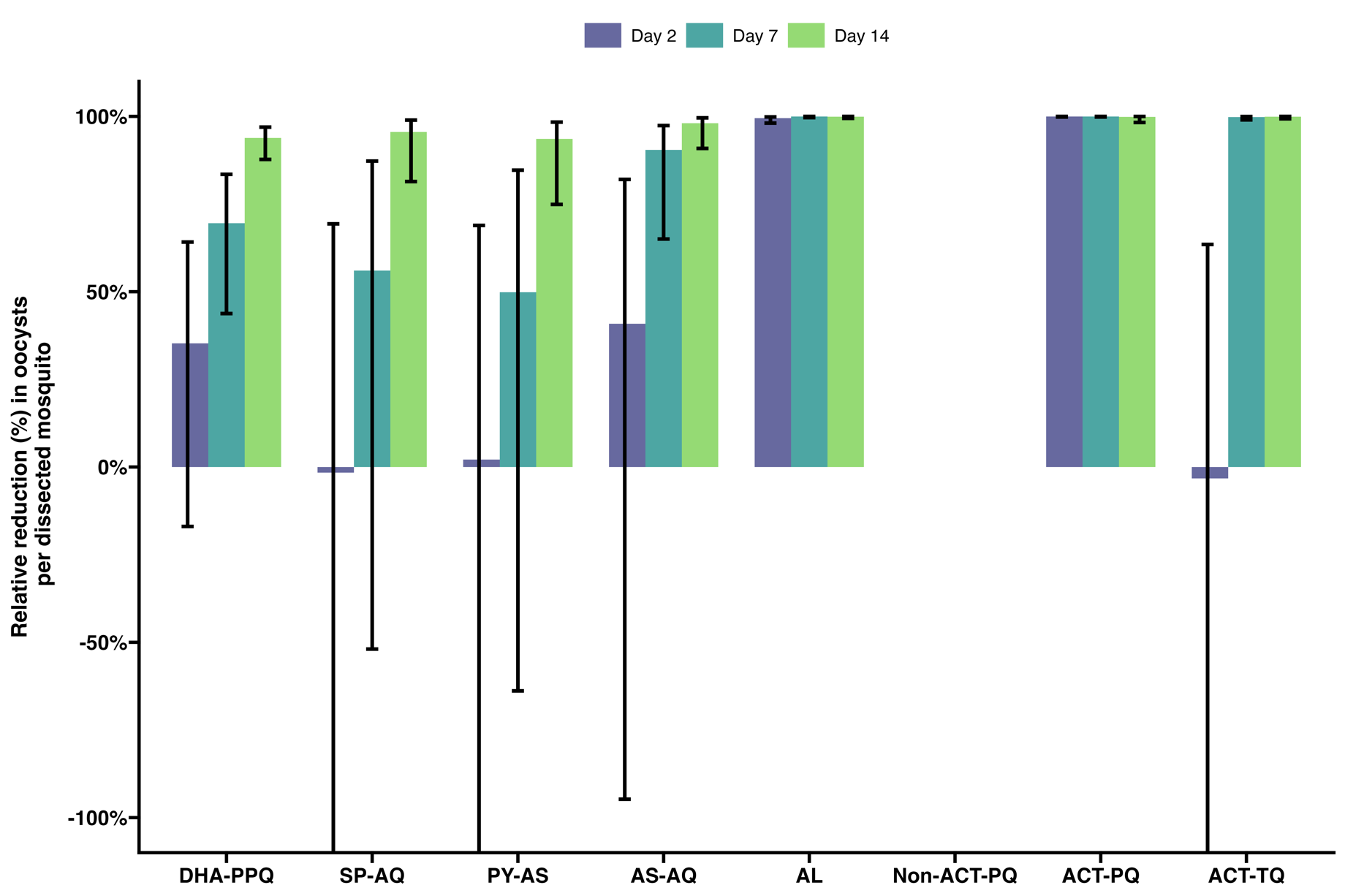

### Supplementary Table 21. Relative reduction in mean oocyst density in dissected mosquitoes at day 2 compared to baseline

| reference | DHA-PPQ | SP-AQ | PY-AS | AS-AQ | AL | ACT-PQ | ACT-TQ |
| --- | --- | --- | --- | --- | --- | --- | --- |
| DHA-PPQ | 35.26% ( -16.96% -64.16%) | p=0.3969 | p=0.4096 | p=0.8643 | p<0.0001 | p<0.0001 | p=0.2845 |
| SP-AQ | p=0.3969 | -1.59% (-236.76% -69.35%) | p=0.9499 | p=0.3799 | p<0.0001 | p<0.0001 | p=0.9763 |
| PY-AS | p=0.4096 | p=0.9499 | 2.13% (-208.08% -68.91%) | p=0.3931 | p<0.0001 | p<0.0001 | p=0.9166 |
| AS-AQ | p=0.8643 | p=0.3799 | p=0.3931 | 40.84% ( -94.75% -82.03%) | p<0.0001 | p<0.0001 | p=0.2982 |
| AL | p<0.0001 | p<0.0001 | p<0.0001 | p<0.0001 | 99.47% (98.09% -99.85%) | p=0.0019 | p<0.0001 |
| ACT-PQ | p<0.0001 | p<0.0001 | p<0.0001 | p<0.0001 | p=0.0019 | 99.97% (99.83% -99.99%) | p<0.0001 |
| ACT-TQ | p=0.2845 | p=0.9763 | p=0.9166 | p=0.2982 | p<0.0001 | p<0.0001 | -3.23% (-191.81% -63.48%) |

Diagonal cells indicate percentage relative reduction at day 2 compared to baseline (95% CI) per treatment category, with corresponding p-value compared to baseline. Other cells in the table contain p-values of between-arm comparisons of the relative reductions.

### Supplementary Table 22. Relative reduction in mean oocyst density in dissected mosquitoes at day 7 compared to baseline

| reference | DHA-PPQ | SP-AQ | PY-AS | AS-AQ | AL | ACT-PQ | ACT-TQ |
| --- | --- | --- | --- | --- | --- | --- | --- |
| DHA-PPQ | 93.88% (87.73% -96.94%) | p=0.6129 | p=0.9439 | p=0.1035 | p<0.0001 | p=0.0019 | p<0.0001 |
| SP-AQ | p=0.6129 | 95.57% (81.42% -98.94%) | p=0.6115 | p=0.3065 | p=0.0003 | p=0.0071 | p=0.0004 |
| PY-AS | p=0.9439 | p=0.6115 | 93.61% (74.89% -98.37%) | p=0.1264 | p<0.0001 | p=0.0024 | p<0.0001 |
| AS-AQ | p=0.1035 | p=0.3065 | p=0.1264 | 98.07% (90.87% -99.59%) | p=0.0041 | p=0.0490 | p=0.0064 |
| AL | p<0.0001 | p=0.0003 | p<0.0001 | p=0.0041 | 99.95% (99.44% -99.99%) | p=0.5060 | p=0.8712 |
| ACT-PQ | p=0.0019 | p=0.0071 | p=0.0024 | p=0.0490 | p=0.5060 | 99.85% (98.27% -99.99%) | p=0.6067 |
| ACT-TQ | p<0.0001 | p=0.0004 | p<0.0001 | p=0.0064 | p=0.8712 | p=0.6067 | 99.93% (99.32% -99.99%) |

Diagonal cells indicate percentage relative reduction at day 7 compared to baseline (95% CI) per treatment category, with corresponding p-value compared to baseline. Other cells in the table contain p-values of between-arm comparisons of the relative reductions.

### Supplementary Table 23. Relative reduction in mean oocyst density in dissected mosquitoes at day 14 compared to baseline

| reference | DHA-PPQ | SP-AQ | PY-AS | AS-AQ | AL | ACT-PQ | ACT-TQ |
| --- | --- | --- | --- | --- | --- | --- | --- |
| DHA-PPQ | 69.52% (43.76% - 83.48%) | p=0.5041 | p=0.3361 | p=0.0468 | p<0.0001 | p<0.0001 | p<0.0001 |
| SP-AQ | p=0.5041 | 56.00% (-51.93% - 87.26%) | p=0.8313 | p=0.0224 | p<0.0001 | p<0.0001 | p<0.0001 |
| PY-AS | p=0.3361 | p=0.8313 | 49.87% (-63.86% - 84.67%) | p=0.0099 | p<0.0001 | p<0.0001 | p<0.0001 |
| AS-AQ | p=0.0468 | p=0.0224 | p=0.0099 | 90.44% (65.03% - 97.39%) | p<0.0001 | p<0.0001 | p<0.0001 |
| AL | p<0.0001 | p<0.0001 | p<0.0001 | p<0.0001 | 99.96% (99.66% -100.00%) | p=0.8497 | p=0.1893 |
| ACT-PQ | p<0.0001 | p<0.0001 | p<0.0001 | p<0.0001 | p=0.8497 | 99.97% (99.83% -100.00%) | p=0.0699 |
| ACT-TQ | p<0.0001 | p<0.0001 | p<0.0001 | p<0.0001 | p=0.1893 | p=0.0699 | 99.81% (99.01% - 99.96%) |

Diagonal cells indicate percentage relative reduction at day 14 compared to baseline (95% CI) per treatment category, with corresponding p-value compared to baseline. Other cells in the table contain p-values of between-arm comparisons of the relative reductions.

### Supplementary Table 24. Hazard ratios for infectivity survival curves (adjusted by baseline PCR gametocyte densities)

| reference | DHA-PPQ | SP-AQ | PY-AS | AS-AQ | AL | Non-ACT-PQ | ACT-PQ | ACT-TQ |
| --- | --- | --- | --- | --- | --- | --- | --- | --- |
| DHA-PPQ |  | 0.91 (0.50- 1.66) p=0.7526 | 1.04 (0.69- 1.56) p=0.8541 | 1.13 (0.74- 1.74) p=0.5667 | 5.26 (3.12- 8.86) p<0.0001 | 8.52 (4.50-16.13) p<0.0001 | 7.93 (5.69-11.04) p<0.0001 | 2.01 (1.26- 3.22) p=0.0037 |
| SP-AQ | 1.10 (0.60- 2.01) p=0.7526 |  | 1.14 (0.76- 1.72) p=0.5179 | 1.25 (0.82- 1.89) p=0.2940 | 5.79 (3.24-10.36) p<0.0001 | 9.38 (4.96-17.75) p<0.0001 | 8.73 (5.13-14.87) p<0.0001 | 2.22 (1.40- 3.50) p=0.0007 |
| PY-AS | 0.96 (0.64- 1.45) p=0.8541 | 0.87 (0.58- 1.32) p=0.5179 |  | 1.09 (1.05- 1.14) p<0.0001 | 5.06 (3.92- 6.53) p<0.0001 | 8.20 (5.54-12.12) p<0.0001 | 7.63 (6.37- 9.14) p<0.0001 | 1.94 (1.75- 2.14) p<0.0001 |
| AS-AQ | 0.88 (0.57- 1.35) p=0.5667 | 0.80 (0.53- 1.21) p=0.2940 | 0.92 (0.88- 0.96) p<0.0001 |  | 4.64 (3.66- 5.88) p<0.0001 | 7.51 (5.26-10.73) p<0.0001 | 6.99 (5.85- 8.36) p<0.0001 | 1.77 (1.66- 1.90) p<0.0001 |
| AL | 0.19 (0.11-0.32) p<0.0001 | 0.17 (0.10-0.31) p<0.0001 | 0.20 (0.15-0.25) p<0.0001 | 0.22 (0.17-0.27) p<0.0001 |  | 1.62 (1.29-2.03) p<0.0001 | 1.51 (1.19-1.91) p=0.0006 | 0.38 (0.32-0.46) p<0.0001 |
| Non-ACT-PQ | 0.12 (0.06-0.22) p<0.0001 | 0.11 (0.06-0.20) p<0.0001 | 0.12 (0.08-0.18) p<0.0001 | 0.13 (0.09-0.19) p<0.0001 | 0.62 (0.49-0.78) p<0.0001 |  | 0.93 (0.64-1.36) p=0.7097 | 0.24 (0.17-0.32) p<0.0001 |
| ACT-PQ | 0.13 (0.09-0.18) p<0.0001 | 0.11 (0.07-0.19) p<0.0001 | 0.13 (0.11-0.16) p<0.0001 | 0.14 (0.12-0.17) p<0.0001 | 0.66 (0.52-0.84) p=0.0006 | 1.07 (0.74-1.57) p=0.7097 |  | 0.25 (0.21-0.30) p<0.0001 |
| ACT-TQ | 0.50 (0.31-0.80) p=0.0037 | 0.45 (0.29-0.71) p=0.0007 | 0.52 (0.47-0.57) p<0.0001 | 0.56 (0.53-0.60) p<0.0001 | 2.61 (2.18-3.14) p<0.0001 | 4.23 (3.10-5.78) p<0.0001 | 3.94 (3.29-4.72) p<0.0001 |  |

Hazard ratios for between-arm comparisons of infectivity clearance, with corresponding 95% CI and p-value.

E.g. Hazard ratio of infectivity clearance for AL compared to DHA-PPQ is 5.26, meaning that clearance is 5.26 times more likely to take place in the AL group compared to the DHA-PPQ group, and this is significantly different (p<0.0001).

### Supplementary Table 25. Hazard ratios for gametocytes by microscopy survival curves

| reference | DHA-PPQ | SP-AQ | PY-AS | AS-AQ | AL | Non-ACT-PQ | ACT-PQ | ACT-TQ |
| --- | --- | --- | --- | --- | --- | --- | --- | --- |
| DHA-PPQ |  | 1.01 (0.60- 1.71) p=0.9598 | 1.35 (0.87- 2.10) p=0.1822 | 1.42 (0.88- 2.28) p=0.1492 | 2.86 (1.73- 4.71) p<0.0001 | 2.23 (1.47- 3.37) p=0.0002 | 6.50 (3.81-11.09) p<0.0001 | 3.67 (2.19- 6.17) p<0.0001 |
| SP-AQ | 0.99 (0.59- 1.66) p=0.9598 |  | 1.33 (0.81- 2.19) p=0.2600 | 1.40 (0.85- 2.31) p=0.1886 | 2.82 (1.77- 4.49) p<0.0001 | 2.20 (1.34- 3.60) p=0.0018 | 6.41 (3.97-10.34) p<0.0001 | 3.62 (2.20- 5.97) p<0.0001 |
| PY-AS | 0.74 (0.48-1.15) p=0.1822 | 0.75 (0.46-1.24) p=0.2600 |  | 1.05 (0.99-1.12) p=0.1146 | 2.12 (1.89-2.37) p<0.0001 | 1.65 (1.55-1.75) p<0.0001 | 4.81 (3.72-6.23) p<0.0001 | 2.72 (2.27-3.26) p<0.0001 |
| AS-AQ | 0.70 (0.44-1.13) p=0.1492 | 0.71 (0.43-1.18) p=0.1886 | 0.95 (0.89-1.01) p=0.1146 |  | 2.02 (1.89-2.15) p<0.0001 | 1.57 (1.43-1.72) p<0.0001 | 4.58 (3.56-5.90) p<0.0001 | 2.59 (2.29-2.93) p<0.0001 |
| AL | 0.35 (0.21-0.58) p<0.0001 | 0.35 (0.22-0.56) p<0.0001 | 0.47 (0.42-0.53) p<0.0001 | 0.50 (0.47-0.53) p<0.0001 |  | 0.78 (0.67-0.90) p=0.0008 | 2.27 (1.75-2.96) p<0.0001 | 1.29 (1.15-1.44) p<0.0001 |
| Non-ACT-PQ | 0.45 (0.30-0.68) p=0.0002 | 0.46 (0.28-0.75) p=0.0018 | 0.61 (0.57-0.64) p<0.0001 | 0.64 (0.58-0.70) p<0.0001 | 1.28 (1.11-1.49) p=0.0008 |  | 2.92 (2.34-3.64) p<0.0001 | 1.65 (1.37-1.98) p<0.0001 |
| ACT-PQ | 0.15 (0.09-0.26) p<0.0001 | 0.16 (0.10-0.25) p<0.0001 | 0.21 (0.16-0.27) p<0.0001 | 0.22 (0.17-0.28) p<0.0001 | 0.44 (0.34-0.57) p<0.0001 | 0.34 (0.28-0.43) p<0.0001 |  | 0.57 (0.44-0.72) p<0.0001 |
| ACT-TQ | 0.27 (0.16-0.46) p<0.0001 | 0.28 (0.17-0.45) p<0.0001 | 0.37 (0.31-0.44) p<0.0001 | 0.39 (0.34-0.44) p<0.0001 | 0.78 (0.70-0.87) p<0.0001 | 0.61 (0.50-0.73) p<0.0001 | 1.77 (1.39-2.25) p<0.0001 |  |

Hazard ratios for between-arm comparisons of microscopical gametocyte clearance, with corresponding 95% CI and p-value.

E.g. Hazard ratio of microscopical gametocyte clearance for AL compared to DHA-PPQ is 2.86, meaning that clearance is 2.86 times more likely to take place in the AL group compared to the DHA-PPQ group, and this is significantly different (p<0.0001).

### Supplementary Table 26. Hazard ratios for gametocytes by PCR survival curves

| reference | DHA-PPQ | SP-AQ | PY-AS | AS-AQ | AL | Non-ACT-PQ | ACT-PQ | ACT-TQ |
| --- | --- | --- | --- | --- | --- | --- | --- | --- |
| DHA-PPQ |  | 0.42 (0.18- 0.96) p=0.0395 | 1.07 (0.73- 1.56) p=0.7219 | 1.27 (0.76- 2.10) p=0.3606 | 2.91 (1.47- 5.76) p=0.0021 | 6.62 (3.88-11.29) p<0.0001 | 24.94 (12.19-51.02) p<0.0001 | 6.01 (3.61- 9.99) p<0.0001 |
| SP-AQ | 2.38 (1.04-5.44) p=0.0395 |  | 2.55 (1.38- 4.71) p=0.0028 | 3.02 (1.23- 7.41) p=0.0161 | 6.93 (2.16- 22.25) p=0.0011 | 15.76 (8.13- 30.56) p<0.0001 | 59.39 (19.90-177.26) p<0.0001 | 14.31 (5.79- 35.32) p<0.0001 |
| PY-AS | 0.93 (0.64- 1.36) p=0.7219 | 0.39 (0.21- 0.72) p=0.0028 |  | 1.18 (0.81- 1.72) p=0.3795 | 2.72 (1.47- 5.04) p=0.0015 | 6.18 (5.00- 7.64) p<0.0001 | 23.29 (13.10-41.41) p<0.0001 | 5.61 (3.85- 8.17) p<0.0001 |
| AS-AQ | 0.79 (0.48- 1.31) p=0.3606 | 0.33 (0.13- 0.81) p=0.0161 | 0.85 (0.58- 1.23) p=0.3795 |  | 2.30 (1.41- 3.74) p=0.0008 | 5.23 (3.97- 6.89) p<0.0001 | 19.69 (13.06-29.70) p<0.0001 | 4.74 (4.53- 4.96) p<0.0001 |
| AL | 0.34 (0.17- 0.68) p=0.0021 | 0.14 (0.04- 0.46) p=0.0011 | 0.37 (0.20- 0.68) p=0.0015 | 0.43 (0.27- 0.71) p=0.0008 |  | 2.27 (1.26- 4.11) p=0.0064 | 8.57 (4.69-15.63) p<0.0001 | 2.06 (1.27- 3.36) p=0.0037 |
| Non-ACT-PQ | 0.15 (0.09-0.26) p<0.0001 | 0.06 (0.03-0.12) p<0.0001 | 0.16 (0.13-0.20) p<0.0001 | 0.19 (0.15-0.25) p<0.0001 | 0.44 (0.24-0.79) p=0.0064 |  | 3.77 (2.26-6.27) p<0.0001 | 0.91 (0.69-1.20) p=0.4959 |
| ACT-PQ | 0.04 (0.02-0.08) p<0.0001 | 0.02 (0.01-0.05) p<0.0001 | 0.04 (0.02-0.08) p<0.0001 | 0.05 (0.03-0.08) p<0.0001 | 0.12 (0.06-0.21) p<0.0001 | 0.27 (0.16-0.44) p<0.0001 |  | 0.24 (0.17-0.35) p<0.0001 |
| ACT-TQ | 0.17 (0.10-0.28) p<0.0001 | 0.07 (0.03-0.17) p<0.0001 | 0.18 (0.12-0.26) p<0.0001 | 0.21 (0.20-0.22) p<0.0001 | 0.48 (0.30-0.79) p=0.0037 | 1.10 (0.83-1.46) p=0.4959 | 4.15 (2.86-6.02) p<0.0001 |  |

Hazard ratios for between-arm comparisons of molecular gametocyte clearance, with corresponding 95% CI and p-value.

E.g. Hazard ratio of molecular gametocyte clearance for AL compared to DHA-PPQ is 2.91, meaning that clearance is 2.91 times more likely to take place in the AL group compared to the DHA-PPQ group, and this is significantly different (p=0.0021).
